# Modeling uncertainty in individual predictions of cognitive functioning for untreated glioma patients using Bayesian regression

**DOI:** 10.1101/2024.10.08.24315003

**Authors:** S.M. Boelders, B. Nicenboim, E. Postma, G.J.M Rutten, K Gehring, L.L. Ong

**Author notes:** Correspondence concerning this article should be addressed to Karin Gehring, PhD. Neurosurgery department, Elisabeth-Tweesteden hospital / Cognitive Neuropsychology, Tilburg University., P. O. Box 90153, Warandelaan 2, Tilburg 5000 LE, The Netherlands.

## Abstract

**Introduction:** Cognitive impairments of patients with a glioma are increasingly considered when making treatment decisions considering a personalized onco-functional balance. Predicting cognitive functioning before surgery can serve as a steppingstone for the clinical goal of predicting cognitive functioning after surgery. However, in a previous study, machine-learning models could not reliably predict cognitive functioning before surgery using a comprehensive set of clinical variables. The current study aims to improve predictions while making the uncertainty in individual predictions explicit.

**Method:** Pre-operative cognitive functioning was predicted for 340 patients with a glioma across eight cognitive tests. This was done using six multivariate Bayesian regression models following a machine-learning approach while using a comprehensive set of clinical variables. Four models included interactions with- or a multilevel structure over histopathological diagnosis. Point-wise predictions were compared using the coefficient of determination (R^2^) and the best-performing model was interpreted.

**Results:** Bayesian models outperformed machine-learning models and benefitted from using shrinkage priors. The R^2^ ranged between 0.3% and 21.5% with a median across tests of 7.2%. Estimated errors of individual prediction were high. The best-performing model allowed parameters to differ across histopathological diagnoses while pulling them toward the population mean.

**Conclusion:** Bayesian models can improve predictions while providing uncertainty estimates for individual predictions. Despite this, the uncertainty in predictions of pre-operative cognitive functioning using the included clinical variables remained high. Consequently, clinicians should not infer cognitive functioning from these variables. Different histopathological diagnoses are best treated as distinct yet related.

**Highlights:**

- Bayesian regression outperformed machine-learning models.
- Predictions were uncertain despite improvements.
- Different histopathological diagnoses are best treated as distinct yet related.

**Importance of the study:** Cognitive impairments of patients with a glioma are increasingly considered when making treatment decisions considering a personalized onco-functional balance. Predicting cognitive functioning before surgery serves as a steppingstone for the clinical goal of predicting cognitive functioning after surgery. The current study is important for two reasons. First, it demonstrates that Bayesian models can improve predictions of pre-operative cognitive functioning over popular machine-learning models. Second, it explicitly shows that individual predictions of pre-operative cognitive functioning based on a comprehensive set of readily available clinical variables included in the current study are uncertain. Consequently, clinicians should not infer cognitive functioning from these variables. Last, it shows that prediction models may benefit a multifaceted view of patients and from treating patients with different histopathological diagnoses as distinct yet related.

## Introduction

Patients with a glioma often suffer from cognitive impairments which are a great burden for both the patients and their caregivers^1^. Cognitive impairments affect quality of life^2^, functional independence^3^, and medical decision-making capacity^4^, and are affected by many variables including variables describing the tumor^3,5–11^, patient characteristics^5,12,13^, medicine use^6,14,15^, and overall measures of health^13,16,17^.

The consideration of cognitive functioning is becoming increasingly important in patient counseling^4,18,19^ and in determining the appropriate treatment in view of a personalized onco-functional balance (i.e. balancing treatment against loss of function)^20^. Ideally, clinicians would be able to employ predictions of cognitive functioning after surgery for individual patients using clinical variables readily available before surgery. Predicting cognitive functioning before surgery can serve as a steppingstone in the process of developing models for the clinical goal of predicting cognitive functioning after surgery. In a previous study, however, we found that machine-learning models could not reliably predict cognitive functioning before surgery while using a comprehensive set of clinical variables^21^.

The ability to obtain reliable predictions is hampered by two sources of uncertainty: aleatoric uncertainty and epistemic uncertainty^22^. Aleatoric uncertainty refers to the inherent randomness in most real-world phenomena and cannot be reduced, even with more data or a better understanding of causal mechanisms. One example of this is the test-retest variability observed in measurements of neuropsychological functioning. In contrast, epistemic uncertainty arises from an incomplete understanding of the causal mechanisms behind the observed data and can be mitigated by improving our understanding or obtaining additional data. One example of this is the incomplete understanding of the mechanisms by which glioma affect cognitive function.

Bayesian models as used in the current study offer two advantages compared to the machine-learning models employed in our previous study. First, they can model the uncertainty in individual predictions^23^. This differs from frequentist models where, by default, we only get point estimates. These uncertainty estimates allow for explicitly describing the amount of uncertainty associated with individual predictions. Moreover, they can be used to assess if an individual prediction can be relied upon. This is important for healthcare applications, especially when using predictions to make important treatment decisions. Such uncertainty estimates allow models to say ‘I don’t know’ instead of being confidently wrong^24^.

Bayesian models obtain such uncertainty estimates by modeling distributions over the model parameters through a process called Markov Chain Monte Carlo (MCMC) sampling. MCMC draws numerous samples from the model parameters based on the training data and prior assumptions made about the model parameters. This process results in an updated distribution of the model parameters, known as the posterior distribution. The posterior distribution of the model parameters consequently informs the distribution over an individual prediction when given a set of predictors. This distribution is called the posterior predictive distribution and can be used to determine a point estimate. Bayesian predictions thus provide a point estimate for each individual patient including a probabilistic distribution describing the probability of the different outcomes. Bayesian models have already been successfully used to describe neurological functions in the field of computational neuropsychology^25^, in research on mental health^26^, and for the clinical application of predictions in healthcare settings^27^. Additionally, their use is becoming easier with user-friendly software packages^28^.

The second advantage of Bayesian models is that they allow for incorporating prior knowledge, not only into the model structure but also into the parameter estimates using priors. These priors represent what we believe to be true about model parameters before having seen any data. Using priors can potentially improve performance compared to not using priors, as is the case with frequentist models^27^. Moreover, shrinkage priors such as horseshoe priors can be set over a collection of parameters to incorporate expectations regarding the magnitude and number of non-zero coefficients priors^29^. Using priors likely benefits model performance when predicting cognitive functioning as sample sizes are typically small relative to the multitude of potential predictors.

The current study predicts pre-operative cognitive functioning for 340 patients with a glioma using six multivariate Bayesian regression models of increasing complexity. This was done while employing a comprehensive set of pre-operatively known predictors that are readily available and have been associated with cognitive functioning in the literature. Importantly, the current study uses the same sample and predictors as our previous study^21^ where cognitive functioning was predicted using popular machine-learning models, allowing us to compare results. We hypothesized that better performance can be achieved with Bayesian models when compared to frequentist models while having the additional benefit of making the uncertainty in individual predictions explicit.

## Method

### Participants

We included 340 patients with grade 2, 3, and 4 gliomas who underwent surgery at the Elisabeth-TweeSteden Hospital, Tilburg, The Netherlands, between 2010 and 2019 and underwent pre-operative cognitive screening as part of clinical care. Patients were not included when they were under 18, had a progressive neurological disease, had a psychiatric or acute neurological disorder within the past two years, or had reduced testability for the neuropsychological testing. For normative purposes, data from healthy Dutch adults were used^30,31^. This study was part of a protocol registered with the Medical Ethics Committee Brabant (file number NW2020-32). This is the same sample as (in part) included in^21,32–36^.

### Interview and cognitive testing

Patients provided informed consent to use their clinical data for research purposes after which a standardized interview was performed. This standardized interview was performed to obtain five of the included predictors; age, sex, and education (the Dutch Verhage scale), and symptoms of anxiety and depression measured using the Dutch translation of the Hospital Anxiety and Depression Scale (HADS)^37^.

All patients performed the computerized CNS Vital Signs (CNS VS)^38^ brief neuropsychological test battery. The psychometric properties of this test battery were shown to be similar to the pen-and-paper tests in patients with various neuropsychiatric disorders and healthy participants ^31,39–41^. Before starting each test, instructions were provided by a well-trained technician (neuropsychologist or neuropsychologist in training). Afterward, the technician reported on the validity of each test within the test battery. Requirements for the validity of the tests included the patient understanding the test, showing sufficient effort, having no vision or motor impairments that affect the task, and the absence of any (external) distractions. Tests that were labeled as invalid were excluded on a test-by-test basis.

### Cognitive test measures and standardization

Test scores were calculated from the CNS VS results according to the formulas presented in Appendix 1, resulting in eight different raw scores. Test scores were scaled such that a higher test score represents a better performance. Scores were converted to socio-demographically adjusted z-scores by correcting for effects of age, sex, and education as found in a sample of normative controls using a multiple regression approach^21,30^. Test scores were standardized relative to those of healthy participants, where test scores for healthy participants were set to have a mean of zero and a standard deviation of one. All preprocessing steps as performed in the current study are visualized in Figure 1 and were the same as done in our previous study^21^.

**Figure 1.**
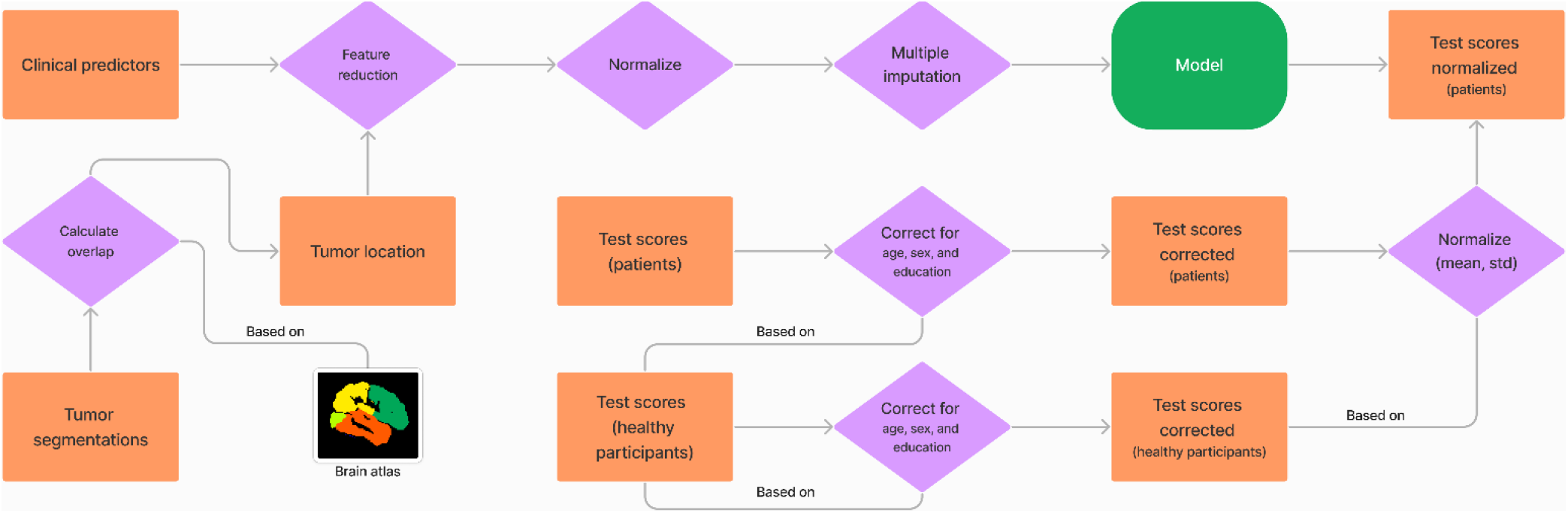
Data preprocessing. Flowchart of the preprocessing steps performed for both the predictors and cognitive test scores. Rectangles represent data, diamonds represent an operation, arrows represent the flow of data, and lines indicate a dependency between an operation and data. The place where the model is fitted is represented as a rounded rectangle.

**Figure 2.**
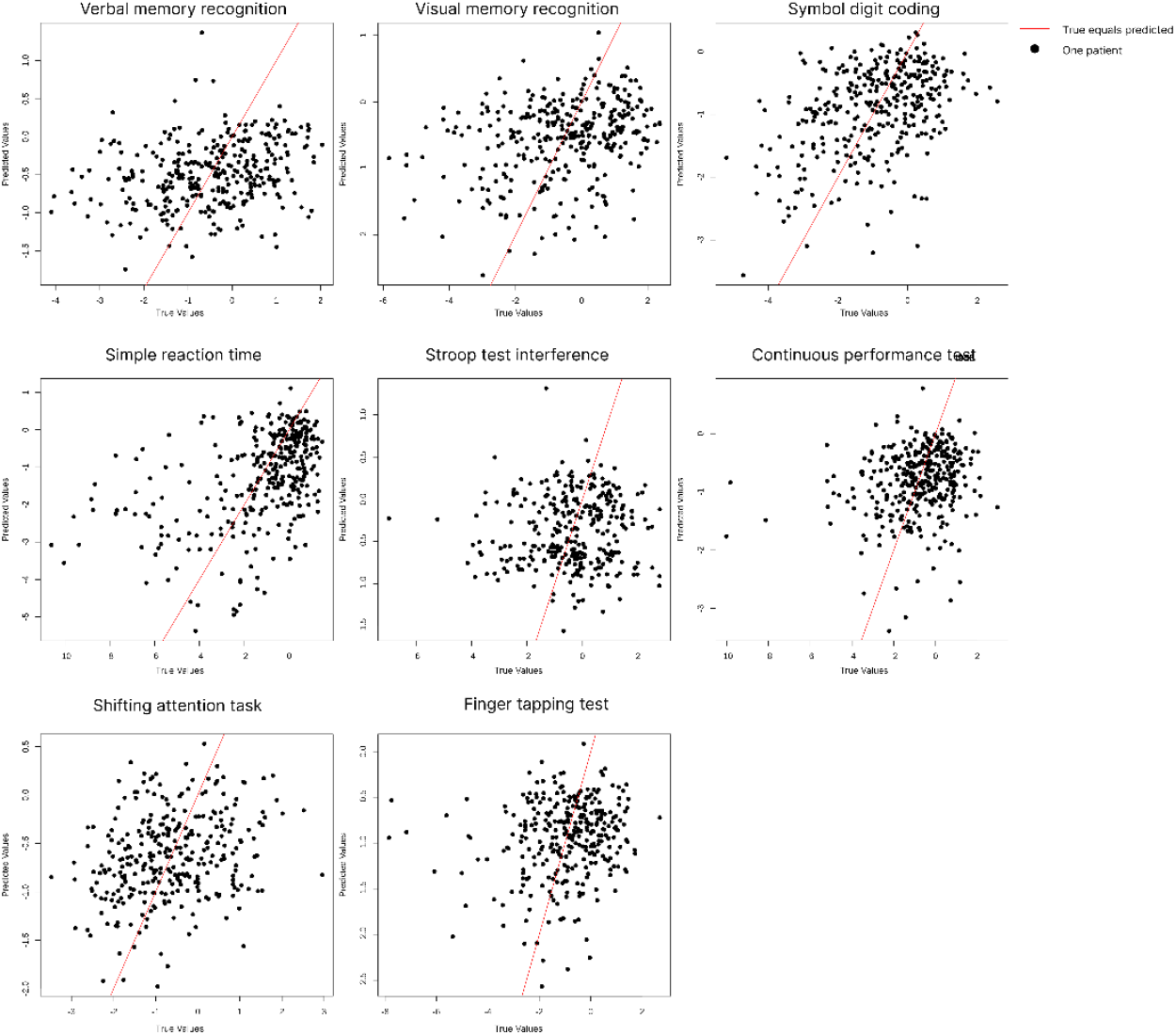
Predicted values versus measured values. Scatter plots of predicted values versus the measured values, individually for each outcome measure. Each dot represents a patient their true and predicted test score for a given test. The red line represents perfect predictions where the predicted value is equal to the measured value (x=y).

### Clinical characteristics

Eight of the included predictors were collected from patients’ electronic medical files. The set of pre-consisted of tumor grade classified according to the WHO guidelines as used at the time of treatment^42,43^, histopathological diagnosis (oligodendroglioma, astrocytoma, or glioblastoma as based on cell origin/molecular markers), IDH mutation status, involved hemisphere, use of antiepileptic drugs, comorbidities, ASA score (the physical status of the patient before surgery^44^), and presenting symptoms. The presenting symptoms were recorded by the neurosurgeon during the first consultation and were later classified into five binary categories indicating the presence or absence of a symptom: behavioral/cognitive problems, language problems, epilepsy/loss of consciousness, motor deficits including paresis, and headache.

Note that for the histopathological diagnosis, IDH mutation status, and tumor grade, the measured values were used while they can only be *estimated* pre-operatively^45^. Moreover, in our clinical practice, the IDH mutation status of patients aged over 55 with a grade 4 glioblastoma is not always tested as the incidence rate of IDH mutant gliomas in this group is very low^46–48^. Therefore, missing IDH mutation statuses for these patients were set to be wild-type. A detailed rationale behind this choice is provided in Appendix 2.

### Tumor volume and location

Two of the included predictors were tumor volume and location. To obtain these predictors, anatomical MRI scans (T1, T1 contrast, T2, FLAIR) were collected when available and registered to MNI space using affine transformation. Tumors were automatically segmented using a convolutional neural network with a U-Net architecture^49,50^ and segmentations were manually validated. Incorrect segmentations were redone semi-automatically. The region making up the tumor was defined as the hyperintense region of the FLAIR scan for low-grade gliomas and the hyperintense region of the T1 contrast for high-grade gliomas. The volume of the tumor was defined as the number of voxels (mm^3^) in the segmentation. Location was defined as the percentage of overlap between the segmentations and the four lobes using individually for each hemisphere. Eight regions were used as higher resolution parcellation likely do not lead to better prediction performance^32^. More details regarding registration and segmentation can be found in Appendix 3.

### Modeling

#### Variable reduction

Even though the models used in the current study can handle large numbers of predictors due to using shrinkage priors (see the next section), too many variables may hinder model convergence, affect accuracy, and result in a large amount of uncertainty. Therefore, the number of variables was reduced such that at least 10% of patients were in a given category, the variance inflation factors were below 0.5, the pairwise correlations between to-be-combined variables were above 0.6, and such that the resulting variables were interpretable. This was the same as done in our previous study where the process is described in detail^21^. Last, all variables used for prediction were normalized to have a mean of zero and a standard deviation of one to ensure all variables contribute equally during model fitting and to aid the interpretation of model parameters.

#### Bayesian regression models

Two versions (a/b) of three Bayesian models (models 1, 2, and 3) were evaluated to predict the eight outcome measures describing cognitive functioning, resulting in a total of six models.

**Model 1** was a multiple multivariate linear regression model (i.e., a model with multiple dependent variables and multiple predictors). A multivariate model was used to allow model parameters for different test scores to be jointly estimated.

**Model 2** was the same as the first while additionally including interaction effects between predictors and the histopathological diagnoses (oligodendroglioma, astrocytoma, or glioblastoma). This was done as predictors of cognitive function, despite being related, may differ across diagnoses^11^.

**Model 3** was also a multiple multivariate linear model. However, this model allowed coefficients to differ between different histopathological diagnoses by using partial pooling. This method models the coefficients as coming from the same distribution, enabling them to vary while also pulling them toward the population average.

All three models were evaluated with (b) and without (a) modeling residual correlations between the test scores which may benefit performance as cognitive test scores are known to be correlated^51^. Models were implemented using the Bayesian Regression Models using Stan (BRMS) package (v2.20.1)^28,52,53^. A formal description of these models and the BRMS syntax are provided in Appendix 4 and 5 respectively.

For these six models, weakly informative priors were used for the coefficients, intercept, residuals and random effects. This was done as determining informative priors was not possible for two reasons. First, previous studies used a variety of neuropsychological tests to measure cognitive functioning which differ in their sensitivity and the cognitive domains they measure. Therefore, coefficients and intercepts may not translate to our tests. Second, we do not expect our model parameters to be independent, making it difficult to determine informative priors. The weakly informative priors provided some information on the range of possible model parameters while letting the data provide most of the evidence^54^. Alongside the weakly informative priors, expectations regarding the number of non-zero coefficients and the magnitude of coefficients were set using horseshoe priors^29,55^. Horseshoe priors restrict the complexity of the potential models by shrinking both the magnitude of coefficients and the number of non-zero coefficients. Details regarding each individual priors are provided in Appendix 6.

Missing cognitive test scores were estimated directly within the Bayesian models while missing predictors were imputed before model fitting using multiple imputation. Thirty different imputed datasets were created using MICE^56^ (v3.16.0), and models were fitted individually on each dataset. Afterward, model parameters were pooled to account for the uncertainty in imputation. The Bayesian Analysis Reporting Guidelines by Kruschke^57^ were followed.

To inspect if the priors correctly modeled our expectations, prior predictive checks were performed for all models. This involves simulating data from the model before observing the training data (i.e., the prior predictive distributions) and comparing it against the measured data. For the best-performing model (defined below), this prior predictive distribution was visualized for each cognitive test. Models were fitted using the Hamiltonian Monte Carlo algorithm in STAN (v2.21)^58^. Documented R (v4.0.4)^59^ code including dummy data is provided as an online supplement.

### Model convergence evaluation

To determine if models covered the full parameter space, the effectiveness of the Markov Chain Monte Carlo (MCMC) sample chains was evaluated using the effective sample size (ESS). To determine if models converged, Rhat values comparing between- and within-chain estimates for model parameters were reported. Finally, warnings provided by BRMS were reported.

The out-of-sample prediction accuracy of point-wise predictions was evaluated using 10-fold cross-validation. Point-wise predictions were defined as the mean of the posterior predictive distribution. Performance was described as the amount of variance explained (R^2^)^60^, and the best-performing model was defined as having the highest median R^2^ across the eight cognitive tests. Normalization and imputation of the predictors were performed within the cross-validation loop to prevent information leakage^61^. All available test scores were considered in model evaluation regardless of whether the score for a different cognitive test was missing for the same patient. Median performances across tests and performances for individual tests were compared across models and against the machine-learning models in our previous study which were evaluated in the same manner^21^. Point estimates resulting from cross-validation were used to determine performance instead of Bayesian estimates of out-of-sample generalizability^62,63^ to allow this comparison.

To assess if the best-performing model was a good fit for our data, the posterior predictive distributions resulting from this model were visualized. This involves simulating data from the model after conditioning it on the measured data and visualizing the resulting distributions (i.e., the posterior predictive distribution) alongside the measured data.

### Sensitivity to selected priors

To check that the priors were only weakly informative, the fitted model parameters and their credibility intervals (i.e., the posterior distributions) were examined for the best-performing model. Moreover, to check that the priors were broad enough, and to differentiate between the effect of shrinkage priors and the weakly informative priors, the performance of the best-performing model was compared against three additional versions of the same model. These were this same model where the horseshoe prior (version c of the best-performing model), all weakly informative priors (version d), or all priors (version e) were replaced with the default priors in BRMS.

### Explaining the performance of Bayesian models

To understand the performance of the simplest Bayesian model (model 1a: the multivariable multiple regression models without residual correlations) and the effect of using multiple imputed datasets, three additional versions of this model were fitted. These were versions of model 1a with uninformative default priors as fitted on only one imputed dataset (model 1c), with uninformative default priors as fitted on all thirty imputed datasets (1d), and as fitted on all thirty imputed datasets with all default priors except the horseshoe priors (models 1e) or the weakly-informative priors without the horseshoe prior (model 1f).

To facilitate the comparison of the Bayesian models with the frequentist machine-learning models in our previous study, two plain frequentist multiple linear regression models were fitted for each outcome measure. The first set of frequentist models (Freq1) were fit using only one imputed dataset while the second set of frequentist models (Freq2) were fitted individually on each of the thirty imputed datasets. These models are highly similar to the multivariable multiple Bayesian regression models without correlation and all default priors (model 1c), except that they were optimized using QR factorization (the default optimizer in the R package ‘stats’) instead of Bayesian optimization. For the frequentist models using multiple imputed datasets, the models were fitted individually for each imputed dataset after which predictions were made using each of the models and averaged to obtain a point estimate^64^.

### Model interpretation

To understand how the best-performing model obtains its predictions, the fitted model parameters including their credibility interval (i.e. posterior distributions) were interpreted. Moreover, to inspect if the model made systematic errors, point-wise out-of-sample predictions of the best-performing model were plotted against the measured values for each cognitive test.

## Results

### Descriptive statistics and variable reduction

Descriptive statistics before variable reduction are provided in Table 1. Five patients had a bilateral tumor and were excluded from further analysis. Tumor grades were combined into low-grade (grade 2) and high-grade (grade 3 + 4), ASA scores were combined into ASA I and ASA II + III, use of antiepileptic drugs was combined with epilepsy/loss of consciousness as a presenting symptom, and HADS anxiety and depression were combined. This was the same as described in our previous study^21^ where this result is more extensively discussed. The resulting set of variables consisted of age, sex, education, tumor size, tumor location (tumor overlap with the four lobes per hemisphere, lateralization, tumor grade (low versus high), histopathological diagnosis (oligodendroglioma, astrocytoma, or glioblastoma), IDH mutation status, presenting symptoms (behavioral/cognitive problems, language problems, motor deficits, and headache), corticosteroid use, use of an antiepileptic drug or epilepsy/loss of consciousness, presence of a comorbidity, ASA score (I versus II + III), and the combined anxiety and depression score.

**Table 1:** Sample characteristics including the predictors and cognitive test scores.

| Table 1: Sample characteristics |  |  |  |  |  |  |  |  |  |
| --- | --- | --- | --- | --- | --- | --- | --- | --- | --- |
| Variable name | count | Mean / % | std | min | 25% | 50% | 75% | max | Missing (%) |
| Age | 340 | 53.21 | 14.34 | 18 | 45 | 55 | 64 | 81 | 0.00% |
| Education | 340 | 5.05 | 1.14 | 1 | 4 | 5 | 6 | 7 | 0.00% |
| Sex (m) | 340 | 65.88% |  |  |  |  |  |  | 0.00% |
| Astrocytoma | 340 | 23.53% |  |  |  |  |  |  | 0.00% |
| Glioblastoma | 340 | 63.24% |  |  |  |  |  |  | 0.00% |
| Oligodendroglioma | 340 | 13.24% |  |  |  |  |  |  | 0.00% |
| WHO grade 2 | 340 | 27.65% |  |  |  |  |  |  | 0.00% |
| WHO grade 3 | 340 | 9.41% |  |  |  |  |  |  | 0.00% |
| WHO grade 4 | 340 | 62.94% |  |  |  |  |  |  | 0.00% |
| IDH1 mutation status (mutant) | 246 | 43.50% |  |  |  |  |  |  | 27.65% |
| Lateralization left | 340 | 41.76% |  |  |  |  |  |  | 0.00% |
| Lateralization right | 340 | 59.71% |  |  |  |  |  |  | 0.00% |
| Frontal Lobe left (mm<sup>3</sup>) | 333 | 8531.99 | 20832.66 | 0 | 0 | 0 | 5936 | 164182 | 2.06% |
| Occipital lobe left (mm<sup>3</sup>) | 333 | 791.27 | 3861.01 | 0 | 0 | 0 | 0 | 33466 | 2.06% |
| Parietal lobe left (mm<sup>3</sup>) | 333 | 1860.60 | 6135.62 | 0 | 0 | 0 | 11 | 42487 | 2.06% |
| Temporal lobe left (mm<sup>3</sup>) | 333 | 4353.59 | 12228.73 | 0 | 0 | 0 | 0 | 74049 | 2.06% |
| Frontal lobe right (mm<sup>3</sup>) | 333 | 9243.12 | 18310.81 | 0 | 0 | 166 | 9564 | 99580 | 2.06% |
| Occipital lobe right (mm<sup>3</sup>) | 333 | 987.62 | 4653.98 | 0 | 0 | 0 | 0 | 43885 | 2.06% |
| Parietal lobe right (mm<sup>3</sup>) | 333 | 5139.08 | 13397.25 | 0 | 0 | 0 | 1081 | 77898 | 2.06% |
| Temporal lobe right (mm<sup>3</sup>) | 333 | 7534.59 | 16717.71 | 0 | 0 | 0 | 2731 | 93323 | 2.06% |
| Tumor size (mm<sup>3</sup>) | 333 | 52553.37 | 43485.95 | 305 | 22153 | 42176 | 71417 | 264510 | 2.06% |
| ASA I | 338 | 44.97% |  |  |  |  |  |  | 0.59% |
| ASA II | 338 | 49.70% |  |  |  |  |  |  | 0.59% |
| ASA III | 338 | 5.33% |  |  |  |  |  |  | 0.59% |
| Comorbidity | 340 | 47.65% |  |  |  |  |  |  | 0.00% |
| Corticosteroid use | 340 | 59.41% |  |  |  |  |  |  | 0.00% |
| Antiepileptic drug use | 340 | 48.82% |  |  |  |  |  |  | 0.00% |
| HADS anxiety | 321 | 6.58 | 4.59 | 0 | 3 | 6 | 10 | 19 | 5.59% |
| HADS depression | 321 | 4.66 | 3.72 | 0 | 2 | 4 | 7 | 17 | 5.59% |
| Presents with attention, executive function, memory, and/or behavioral problems | 340 | 22.35% |  |  |  |  |  |  | 0.00% |
| Presents with language problems | 340 | 15.29% |  |  |  |  |  |  | 0.00% |
| Presents with loss of consciousness | 340 | 42.94% |  |  |  |  |  |  | 0.00% |
| Presents with motor deficits | 340 | 23.24% |  |  |  |  |  |  | 0.00% |
| Presents with headache | 340 | 23.82% |  |  |  |  |  |  | 0.00% |
| **Cognitive test scores** |  |  |  |  |  |  |  |  |  |
| Verbal memory recognition | 323 | -0.48 | 1.22 | -3.24 | -1.31 | -0.35 | 0.42 | 2.04 | 5.00% |
| Visual memory recognition | 340 | -0.47 | 1.58 | -3.94 | -1.46 | -0.32 | 0.77 | 2.36 | 0.00% |
| Finger tapping test | 315 | -0.89 | 1.37 | -4.28 | -1.73 | -0.74 | 0.10 | 2.68 | 7.35% |
| Symbol digit coding | 340 | -0.85 | 1.43 | -3.82 | -1.79 | -0.75 | 0.20 | 2.36 | 0.00% |
| Simple reaction time | 334 | -1.25 | 2.21 | -7.68 | -2.07 | -0.51 | 0.30 | 1.47 | 1.76% |
| Stroop interference | 315 | -0.35 | 1.40 | -3.64 | -1.08 | -0.26 | 0.66 | 2.79 | 7.35% |
| Shifting attention task | 315 | -0.61 | 1.09 | -2.35 | -1.45 | -0.71 | 0.17 | 2.96 | 7.35% |
| Continuous performance test | 340 | -0.75 | 1.47 | -4.25 | -1.61 | -0.61 | 0.32 | 2.15 | 0.00% |

The prior predictive check for the best-performing model (model 3b, see below) is reported in Appendix 7. The prior predictive checks for all other models were highly similar. For all models, the prior predictive check showed that the simulated data (depicted in light blue) matches the true data (depicted in dark blue) for a small number of draws from the model parameters and training data. All other simulated data covered a wide but reasonable range of outcomes (with a mean of 0 and tails reaching up to around -30 and 30). This indicates that the selected priors are weakly informative and allow for the model to describe our measured data.

### Model convergence and evaluation

There were no warnings reported by BRMS. All models had a Rhat of at most 1.003, indicating good convergence. Models effectively sampled the parameter space with ESS values of at least 84413 and 51130 for the bulk and tail of the distribution respectively.

The performance of all models is displayed in Table 2. The best performance was obtained by model 3a (partial pooling without residual correlations), with a median amount variance explained of 7.2%. The amount of variance explained ranged between 0.3% for the Stroop interference ratio to 21.5% for the measure of simple reaction time. This model obtained the best performance compared to the other models for the measures of verbal memory recognition, visual memory recognition, simple reaction time, continuous performance, and the finger-tapping task. For all other measures, this model (3a) obtained close to the best performance with the amount of variance differing at most 0.5% of variance for the symbol digit coding test.

**Table 2:**
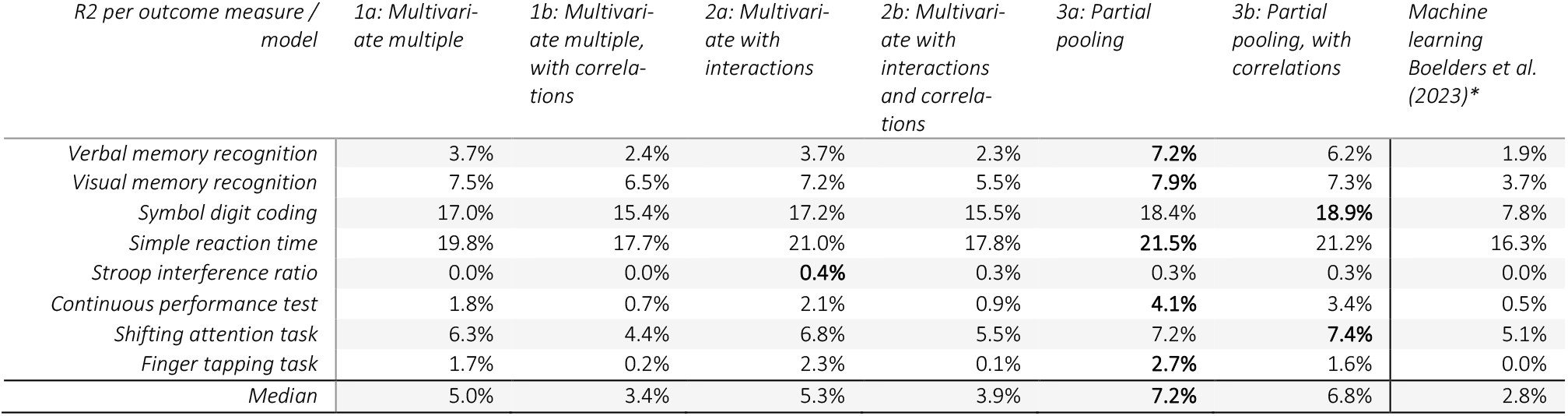
Prediction results. Amount of variance explained (%) per outcome measure and model as determined using 10-fold cross-validation. For reference, the amount of variance explained by the best-performing machine learning model per outcome measure as found in Boelders et al. (2023) is reported. The best result per outcome measure is displayed in bold. *Negative performances were set to zero.

The median performance of all Bayesian models was higher than the median performance of the best-performing machine learning models in our previous study^21^ (Table 2). The best-performing Bayesian model (model 3a) explained an additional 4.4% of variance (median) when compared to the best-performing machine learning models. Moreover, the performance of the best-performing Bayesian model was higher than the machine learning models for all individual cognitive tests with increases in performance ranging from 0.3% of the variance for the Stroop interference ratio to 10.6% of the variance for the symbol digit coding test.

The simplest model, the multiple multivariate regression model (model 1a) had a median explained variance of 5.0%. Adding residual correlations decreased this to 3.4% of variance explained (model 1b). Modeling interactions with histopathological diagnosis (model 2a) increased median performance to 5.3%. Adding residual correlations to this model decreased performance to 3.9% of variance explained. Pooling the coefficients and intercepts resulted in the best models with an explained variance of 7.2% (model 3a), which decreased to 6.8% when modeling residual correlations (model 3b). Thus, conditioning coefficients and intercepts on histopathological diagnosis, and allowing coefficients to differ based on the histopathological diagnosis improved model performance, while modeling residual correlations decreased performance.

### Sensitivity to selected priors

The model parameters of the best-performing model after model fitting (i.e. posterior distributions) (Appendix 8) were within the specified priors, indicating that the priors were broad enough. To test the influence of the selected priors, results for models 3a with (partly) uninformative priors are presented in Table 3 (models 3c, 3d, 3e).

**Table 3:** Prediction results for the prior sensitivity tests and frequentist models. Amount of variance explained (%) per outcome measure and model as determined using 10-fold cross-validation. Results describe the models fitted as sensitivity checks (models 3c-3d) for the best-performing model (3b), the frequentist models (Freq1-2), and the results for model 1 with (partly) default priors (models 1c-1e).

| *R2 per outcome measure \ model* | *3c: Partial pooling, weakly informative, no horseshoe* | *3d: Partial pooling, default, and horseshoe* | *3e: Partial pooling, default priors* | *Freq1: Frequentist regression, one imputed dataset* | *Freq2: Frequentist regression, multiple imputed datasets* | *1c: Default priors, one imputed dataset* | *1d: Default priors* | *1e: Default and horseshoe* | *1f: Weakly informative priors, no horseshoe* |
| --- | --- | --- | --- | --- | --- | --- | --- | --- | --- |
| *Verbal memory recognition* | 6.6% | 7.5% | 7.3% | 0.2% | 0.0% | 3.9% | 3.9% | 3.7% | 3.6% |
| *Visual memory recognition* | 5.5% | 7.1% | 4.9% | 0.0% | 0.1% | 4.5% | 4.6% | 7.6% | 4.9% |
| *Symbol digit coding* | 16.8% | 18.2% | 16.7% | 0.0% | 8.5% | 15.5% | 15.7% | 17.0% | 16.4% |
| *Simple reaction time* | 17.7% | 20.2% | 16.5% | 0.2% | 3.0% | 16.0% | 16.0% | 19.6% | 15.8% |
| *Stroop interference ratio* | 0.0% | 0.3% | 0.0% | 0.5% | 0.2% | 0.0% | 0.0% | 0.0% | 0.0% |
| *Continuous performance test* | 3.4% | 4.0% | 3.1% | 0.1% | 0.7% | 2.8% | 2.9% | 1.8% | 3.0% |
| *Shifting attention task* | 6.0% | 6.7% | 6.0% | 1.2% | 2.5% | 5.5% | 5.4% | 6.3% | 5.3% |
| *Finger tapping task* | 4.9% | 2.5% | 4.4% | 0.0% | 0.7% | 4.5% | 4.4% | 1.8% | 4.4% |
| *Median* | 5.8% | 6.9% | 5.4% | 0.1% | 0.8% | 4.5% | 4.5% | 5.0% | 4.6% |

Removing the horseshoe prior from the best-performing model resulted in a median explained variance of 5.8% (model 3c) relative to 7.2% resulting from the best-performing (3a). This shows the value of using the horseshoe prior. The variant of the best-performing model with all default priors except for the horseshoe prior resulted in a median explained variance of 6.9% (model 3d), which is only a 0.3 percentage point decrease when compared to the best-performing model. This shows that the weakly informative priors only had a small influence on model performance. Finally, the model with only default priors (model 3e) resulted in a median explained variance of 5.4%. This is 1.4 percentage points lower when compared to the best-performing model, showing that the selected priors were suitable. Moreover, this is 0.4 percentage points lower compared to model 3c which uses weakly informative priors and no horseshoe prior, again showing that the weakly informative only had a small influence on performance.

### Explaining the performance of Bayesian models

To explain the performance of the Bayesian models, the simplest Bayesian (model 1a) was compared to a variant with uninformative priors that was fitted on only one imputed dataset (model 1c), variants of this model with (partly) uninformative priors fitted on the thirty imputed datasets (model 1d, 1e, and 1f), and frequentist models fitted on one (model Freq1) or thirty imputed datasets (model Freq2). Results for these additional models are displayed in Table 3.

Removing the horseshoe prior from model 1a resulted in a median performance of 4.6% (model 1f) of variance (versus 5.0%, model 1a). Removing the weakly informative priors but not the horseshoe prior resulted in a median performance of 5.0% (model 1e). Using all default priors resulted in a median performance of 4.5% (model 1d). These results again show the limited effect of the semi-informative priors while also demonstrating the benefits of using the horseshoe prior. These results are in line with the sensitivity checks for model 3b.

The frequentist regression model using one imputed dataset (Freq1) had a median performance of 0.1% of variance, which increased to 0.8% when using thirty imputed datasets (Freq2). Comparing the frequentist regression as evaluated on one imputed dataset (Freq1) to its Bayesian equivalent as fitted on one imputed dataset (model 1c), performance increases from a median explained variance of 0.1% to 4.5%. When comparing the frequentist model using thirty imputed datasets (Freq2) to its Bayesian equivalent using thirty imputed datasets and default priors (model 1d), performance increased from 0.8% to 4.5% of variance. These results show that Bayesian optimization when using default priors resulted in better predictions when compared to QR factorization (frequentist). Notably, the variants of model 1 with default priors as fitted on one imputed dataset (model 1c) or averaged across the thirty imputed datasets (model 1a) performed roughly equivalent with a median of 4.5% of variance explained. This shows that averaging over multiple datasets had little effect on performance for Bayesian models, contrary to the frequentist models.

### Model interpretation

To evaluate if the distributions of the simulated data as obtained after model fitting match the observed data, the posterior predictive check for the best-performing model (model 3a) is visualized in Appendix 9, individually for each cognitive test. These checks show that the simulated data (depicted in light blue) matches the true data (depicted in dark blue) for most of the draws from the model parameters and training data. This indicates that the model was able to adequately describe the observed data. For most outcome measures, and especially the measure of simple reaction time, the model did not fully capture the skewness in the cognitive test data.

To interpret the relationships captured by the best-performing model, we refer back to the parameter estimates after model fitting (i.e., the posterior distributions) as visualized in Appendix 8. It is important to note that the relationships identified by the model are purely descriptive of how the model obtains its predictions as we used a machine-learning approach.

Five things can be observed from the model parameters. First, the model relied on most of the predictors. This can be seen from most coefficients (Appendix 8A) being comparable in magnitude with a median magnitude of 0.005 [95% CI: 0.000, 0.025] and only a few coefficients being close to 0 (23 out of 192 coefficients smaller than 0.001). Second, the model did not rely strongly on any particular variable. This can be seen from the three most influential variables being small relative to the variance in the test scores. More specifically, the largest coefficients, which were all found when predicting simple reaction time, were tumor volume with a median estimated coefficient of -0.048 [95% CI: -0.57, 0.08], a tumor located in the right frontal lobe with a median of coefficient -0.047 [95% CI: -0.46, 0.06], and a tumor in the left frontal lobe with a median coefficient of 0.38 [95% CI: -0.06, 0.40]. These coefficients are low relative to the variance in the test scores which was 2.21 for simple reaction time (Table 1). Third, the model was uncertain in its estimation of the coefficients. This can be seen from the credibility interval for most coefficients being wide relative to their magnitude, as exemplified by the largest coefficients provided previously. Fourth, the expected variability in measurements of cognitive functioning (Appendix 8C) was high relative to the standard deviation of the data (see Table 1). More specifically, the expected variability ranged between 1.02 [95% CI: 0.94, 1.11] for the shifting attention task and 2.00 [95% CI: 1.83, 2.15] for simple reaction time, relative to the variance in test scores which ranged between 1.09 for the shifting attention task and 2.21 for the measure of simple reaction time. Fifth, the coefficients showed substantial variability between patients with different histological diagnoses. This can be seen from the average variability of coefficients between groups (i.e. standard deviation of the group-level coefficients) being 0.206 [95% CI: 0.136, 0.402], while the median magnitude of coefficients was only 0.005 [95% CI: 0.000, 0.025] as previously described.

To ensure that there were no systematic errors, out-of-sample predictions were plotted against the true measured values in Figure 1. The points generally cluster around the red line, suggesting that the model does not exhibit systematic errors for most patients. The largest errors occur when patients score exceptionally poorly on a given test (low value on the x-axis). In these cases, the model tends to overestimate performance, as indicated by points appearing in the upper-left region of the plots.

## Discussion

The current study showed that Bayesian regression models resulted in better predictions of pre-operative cognitive functioning for individual patients when compared to the machine-learning models described in our previous study^21^. Despite the performance improvement, the amount of variance explained remained low (ranging between 0.3% and 21.5% with a median of 7.2%). Moreover, models explicitly showed that the estimated error associated with individual predictions were large. These results show that it is not yet possible to make certain inferences about pre-operative cognitive functioning using the included clinical variables.

All Bayesian models performed better than the best-performing machine learning models in our previous study, including the simplest multivariate multiple Bayesian regression model with a shrinkage prior (model 1a). This model is somewhat similar to the ElasticNet model which was amongst the best-performing machine learning models. Moreover, this included the Bayesian model that used default priors, did not model residual correlations, and was fitted on only one imputed dataset (model 1c). This model also was highly similar to and outperformed the frequentist multiple regression models with one outcome each (Freq1) as fitted in the current study. These results indicate that the increase in performance may, in part, be due to Bayesian estimation or the remaining information in the default priors for the intercept and standard error as used by BRMS.

Prediction performance increased when allowing coefficients and intercepts to differ between histopathological diagnoses (model 2). This indicates that coefficients and intercepts are best treated as different across histopathological diagnoses when considering prediction performance, which is in line with previous studies showing that the effects of predictors may differ across different types of tumors^11^. Performance increased further when pulling the coefficients and intercepts toward the average across the different histopathological diagnoses (model 3). This indicates that at least a subset of coefficients and intercepts are related across these groups.

Using weakly informative priors instead of the default priors was of little influence on model performance. This finding is unsurprising given that the weakly informative priors still provide little information to the model. Using the horseshoe prior resulted in better performance when compared to not using horseshoe priors. This indicates that the assumptions regarding the magnitude of coefficients and the number of non-zero coefficients were appropriate. Inspecting the coefficients of the best-performing model showed that most variables contributed weakly to the prediction. The finding that models using a horseshoe prior performed best and that the resulting models relied on most variables without relying strongly on any specific variable indicate that a multifaceted view of patients is necessary to obtain the best possible prediction, which is in line with our previous study^21^.

All models except for the models without interaction effects, partial pooling, or residual correlation (models 1a and 2a) shared some parameter estimates across cognitive test scores. This allowed these models to use more information by estimating missing test performance during model fitting. Despite this, adding residual correlations between outcome measures (models 1b, 2b, and 3b) decreased model performance. This may be explained either by the added number of parameters to estimate or due to the estimations of missing test scores being inaccurate and uncertain like the predictions themself.

Averaging models over multiple imputed datasets to account for uncertainty in imputations improved the performance of frequentist models, but had no effect on the performance of the Bayesian models. This difference can likely be attributed to the Bayesian model already performing better when not using multiple imputed datasets compared to frequentist models, leaving less room for model performance to increase. The limited effect of using multiple imputed datasets for both the Bayesian and frequentist models can likely be explained by the large number of relatively weak relationships found by the models. These weak relationships cause the influence of any specific (imputed) variable to be limited.

Three limitations of the current sample should be mentioned. First, the current sample was collected as part of routine clinical care. Therefore, it only included patients who were suitable for neuropsychological testing and tumor resection. Consequently, the current study did not include patients with, amongst others, patients in need of immediate surgical intervention or patients with significant motor impairments or severe cognitive limitations. This may have limited the variance in cognitive functioning. Second, a brief computerized test battery was used which may be somewhat reliant on processing speed^40^, and which does not measure language function, memory free recall, or visuoconstructive abilities. Therefore, the current study does not rule out that there are other cognitive domains for which predictions can be made with better performance. However, more comprehensive cognitive evaluations are not typically conducted during the presurgical clinical care of patients with a brain tumor. Last, it is important to note that only readily obtainable clinical variables were considered. Therefore, better and more certain predictions may be obtained by using additional information such as measures of structural^65^ and functional^66–68^ connectivity or additional molecular markers^5^.

We believe the current study is highly important as it is the first to explicitly show that individual predictions of pre-operative cognitive functioning for patients with a glioma based on the comprehensive set of clinical predictors included in the current study are uncertain. Therefore, clinicians should not infer cognitive functioning from these predictors. Moreover, our results show that Bayesian models can result in better predictions when compared to popular machine-learning models. Therefore, we hope the current study stimulates the use of Bayesian models, especially when the certainty of individual predictions is important and when sample sizes are small.

Future studies should collect larger multimodal cross-center datasets to allow for more accurate and more certain predictions and for models to generalize across centers. Moreover, to obtain guarantees regarding the certainty of predictions, methods such as conformal prediction^69,70^ or simulation-based calibration^71^ can be used to ensure that the credible intervals are trustworthy for new data. Last, future work may be aimed at predicting cognitive function after surgery using variables available before surgery and can consider predicting outcomes that are more relevant to patients’ daily functioning such as being able to take care of children.

## Conclusion

Bayesian models resulted in better predictions than machine learning models while having the additional benefit of providing estimates of uncertainty for individual predictions. Despite the performance improvements, individual predictions were uncertain. Therefore, we conclude that it is not yet possible to infer the pre-operative cognitive functioning of individual patients from the comprehensive set of clinical variables used in the current study. Results further show that the weakly informative priors only had a small positive impact on performance while shrinkage priors had a larger positive effect. The best-performing model relied on many variables without relying strongly on any particular variable, indicating that a holistic view of a patient likely is necessary to make the best predictions. Results further show that patients with different histopathological diagnoses are best treated as different yet related. Future studies should consider using Bayesian models when datasets are small and when estimates of uncertainty in individual predictions are important. Moreover, future studies should collect larger multimodal cross-center datasets to obtain more reliable predictions. Finally, future studies should move towards predicting cognitive functioning after surgery for models to be useful in clinical practice.

## Supporting information

Example data

Random effects

Bayesian reporting checklist

## Authorship

Experimental design: (SB, KG, BN, LLO, EP), acquisition: (KG), analysis: (SB), interpretation: (SB, KG, BN, LLO, EP, GR). All authors have been involved in the writing of the manuscript and approved the final version.

## Data availability

Data described in this work is not publicly available to protect the privacy of patients. All code used in this study is available as supplementary material.

