## Appendix for "Modeling uncertainty in individual predictions of cognitive functioning for untreated glioma patients using Bayesian regression"

### Appendix 1: Formulas used to obtain the test scores

| Name | Description | Formula |
|---|---|---|
| Verbal memory recognition | Memory recognition for words. Fifteen words are presented one at a time. The subject identified the presented words amongst new words. The immediate condition is at the beginning of the test battery and the delayed condition is at the end. | Number of items correct (immediate and delayed recall) |
| Visual memory recognition | Memory recognition for abstract images. Fifteen images are presented one at a time. Subjects identified the presented images amongst new images. The immediate condition is at the beginning of the test battery and the delayed condition is at the end. | Number of items correct (immediate and delayed recall) |
| Symbol digit coding | Eight symbols are presented on the screen with a corresponding number. Given a row of eight randomly ordered symbols, the participant is asked to provide the matching number for two minutes straight. | Correct responses - incorrect responses |
| Simple reaction time | The subject presses the space bar when a word is presented. | Average reaction time |
| Stroop test interference | The subject presses the space bar if a word is presented and the color of the word does/or does not match its semantic meaning for the congruent and incongruent trials respectively | (Average reaction time on correct responses for the incongruent trials - Average reaction time on correct responses for the congruent trials) / Average reaction time on correct responses for the congruent trials |
| Shifting attention task | A red circle and a blue square are presented on the screen. Given a third shape, the participant needs to match the shape either by color or shape. | Correct responses - incorrect responses |
| Continuous performance test | The subject responds to a target letter amongst distractors for 5 minutes straight. | Average reaction time to the target letter |
| Finger tapping test | The participant presses the spacebar as often as possible within ten seconds. This task is performed three times for each hand. | The average number of presses across left and right trails |

*Caption: Formulas used to obtain the cognitive domains*

### Appendix 2: Imputing IDH mutation status

In the current study, imputation of predictors is performed using MICE (Multiple Imputation by Chained Equations), where each missing variable is estimated based on other variables. For patients aged 55 or higher with a grade IV glioblastoma, missing values on IDH mutation status were imputed based on the literature instead of using multiple imputation. This was done as for this subset of patients, the IDH mutation status is known to be wildtype in 96% of cases[1]. Not imputing these values based on prior knowledge implies that the iterative imputer must learn how to predict IDH status for this subset of patients. The imputer would have to learn how to impute these values from the relatively small number of patients age 55 or higher with a grade 4 glioblastoma patients for whom IDH status is available. Imputing these values using the iterative imputer, therefore, likely would have resulted in less accurate estimations of IDH status when compared to imputing these values based on the literature. The more accurate the predictions of IDH status are, the more likely it is that we can accurately predict cognitive functioning. Therefore, we opted to impute these values based on clinical knowledge.

Note that the goal of imputation differs between predictive modeling and explanatory modeling. In predictive modeling, imputation is done with the goal of obtaining the highest model performance possible. For explanatory modeling, imputation is done with to goal of being able to use participants with missing values without biassing the results of statistical tests.

### Appendix 3: Segmentation models

All anatomical MRI (T1, T1 contrast, T2, Flair) scans were registered to MNI space using affine transformation. Registration was performed using Regaladin[2] from the NiftyReg package which has been shown to perform well for patients with a primary brain tumor[3]. Skull stripping was performed using HD-BET which is designed to be robust to a variety of different lesions[4]. Tumor volume was defined as the FLAIR-enhancing part for low-grade gliomas and the T1 contrast-enhancing part for high-grade gliomas.

Two different models were used for segmentation and the best segmentation was selected manually for each patient. Models used were nnU-Net[5] as trained on T1, T1c, T2, and Flair scans from the BraTS dataset or subsets thereof[6,7] and AGU-Net as available in the Radionics tool using T1c images for high-grade gliomas and FLAIR images for low-grade gliomas[8]. All automatic segmentations were manually validated and incorrect segmentations were redone semi-automatically using the snake tool in ITK-Snap[9].

### Appendix 4: Formal model description

More specifically, for patients $n \in \mathrm{N}$, we have a response matrix $Y$ of shape $8\ x\ |N|$ representing the eight test scores, a matrix $X_p$ of shape $24$ x $|N|$ representing the predictors, and a contrast coding matrix $X_g$ of shape $2\ x\ |N|$ representing the histopathological diagnosis. Here, $X_g\ \cup\ X_p\ =\ X$ and $X_p\ \cap\ X_g =\ \emptyset$. Now, given a vector for the intercepts $\alpha$ of length eight, a fixed effect coefficients matrix $\beta_p$ of shape $24\ x\ 8$, a coefficients matrix for the histopathological diagnosis $\beta_{\mathrm{g}}$ of shape $2\ x\ 8$, and a covariance matrix of the residuals $\Sigma$ with shape $8$ x $8$, we define the multivariate multiple linear regression model as:

$$Model\ 1: Y \sim MVN\left(\alpha + X_g^T \beta_g^T\ +\ X_p^T\ \beta_p, \Sigma\right)$$

Next, given a coefficient matrix for the interaction between histopathological diagnosis and the other predictors $\beta_{\mathrm{interaction}}$ of shape 48 x 8, we define the model including interaction effects with histopathological diagnosis as:

$$\mathrm{M}odel\ 2: Y \sim MVN\left(\alpha +\ X_g^T \beta_{\mathrm{g}}\ +\ X_p^T \beta_p + (\mathrm{X}_g \odot X_p)^T \beta_{\mathrm{interaction}}, \Sigma\right)$$

Finally, given a matrix $u_g$ of shape $24\ x\ 8$ representing the random effects specific to group *g*, a vector $\mu_{\alpha g}$ of length eight representing the random intercepts specific to group *g,* and the covariance matrices $\Sigma_\mu$ for the random effects $\mu_{\mathrm{g}}, \mu_{\alpha\mathrm{g}}$ with shape $25$ x $25$ individually for each $\mathrm{y}$, we define the partial pooling model as:

$$Model\ 3: Y \sim MVN\left(\alpha + \mu_{\alpha\mathrm{g}}\ +\ X_p^T(\beta\ + \mu_{\mathrm{g}}), \Sigma\right)$$

$$\text{where } (\mu_{\mathrm{g}}, \mu_{\alpha\mathrm{g}}) \sim N\left(0, \Sigma_\mu\right)$$

Each model was evaluated (b) with residual correlations between outcome measures and (a) without residual correlations between outcome measures. In other words, for models with residual correlations (b), $\Sigma$ is a full covariance matrix, while for models without residual correlations (a), $\Sigma$ is a diagonal matrix.

### Appendix 5: BRMS formulas

Given patients $n \in \mathrm{N}$, we have a response matrix $Y$ of shape $8\ x\ |N|$ representing the eight test scores, a matrix $X_p$ of shape $24\ x\ |N|$ representing the predictors, a contrast coding matrix $X_g$ of shape $2\ x\ |N|$ representing the histopathological diagnosis (obtained $as.factor(g)$ where $g$ is a vector of length |N| with three categories). Here, $X_g \cup X_p = X$ and $X_p \cap X_g = \emptyset$. Then we have the following BRMS syntax:

1. A multiple multivariate regression model
   a. With residual correlations between test scores

   $$mvbind(Y) \mid mi() \sim X + set\_rescor(TRUE)$$

   b. Without residual correlations between test scores.

   $$mvbind(Y) \mid mi() \sim X + set\_rescor(FALSE)$$

2. A multilevel multiple multivariate regression model including interaction effects with the histopathological diagnosis of the tumor
   a. With residual correlations between test scores

   $$mvbind(Y) \mid mi() \sim X_p * as.factor(g) + set\_rescor(TRUE)$$

   b. Without residual correlations between test scores.

   $$mvbind(Y) \mid mi() \sim X_p * as.factor(g) + set\_rescor(FALSE)$$

3. A multilevel multiple multivariate regression model conditioned on the histopathological diagnosis with partial pooling of coefficients and intercepts over the different diagnoses
   a. With residual correlations between test scores

   $$mvbind(Y) \mid mi() \sim X_p + (1 + X \mid g) + set\_rescor(TRUE)$$

   b. Without residual correlations between test scores.

   $$mvbind(Y) \mid mi() \sim X_p + (1 + X \mid g) + set\_rescor(FALSE)$$

### Appendix 6: Prior specification

The priors for the intercepts were set to be normally distributed with a mean of 0 and a standard deviation of 3 $\left(\alpha \sim \mathcal{N}(0,3)\right)$. This was done as cognitive scores were scaled relative to healthy participants which have a mean of 0 and a standard deviation of 1. A standard deviation of 3 was chosen as the sample of patients had a standard deviation in test scores ranging between 1.1 and 2.2 depending on the test (see the descriptive statistics in Table 1).

The output distribution was modeled as a multivariate normal distribution as test scores are largely normally distributed and this allows for modeling residual correlations between outcome measures. The prior for the standard error of individual predictions (the diagonal in $\Sigma$) was modeled as a half-normal distribution with a location of 0 and a scale of 3 ($\sigma_\epsilon \sim \mathcal{N}_+(0,3)$). A scale of 3 was chosen as it is larger than the sample's standard deviation, thus ensuring that model certainty comes from the data and not from the prior.

For the models including residual correlations between outcome measures (models 1b, 2b, and 3b), the prior for the covariance matrix $\Sigma$ was set to be the Lewandowski-Kurowicka-Joe distribution with the value for the shape set to 6 to expect most values to be around zero while also allowing for moderate correlations of around 0.5.

For the partial pooling models (models 3a, and 3b), the prior for the standard deviation between parameter estimates across groups (the diagonal of $\Sigma_u$) was set to a half normal with a location of 0 and a scale of 0.3 ($\sigma_u \sim \mathcal{N}_+(0,0.3)$). This was done as we expect coefficients to be small for most parameters, leading to even smaller standard deviations in parameter estimates.

The horseshoe priors were configured to expect 44% of the coefficients to be non-zero. This was done for two reasons. First, based on our previous study, models are expected to rely on many variables without depending strongly on any specific variable[10]. Second, the current study includes a relatively large number of predictors (n=26) relative to the number of patients (n=340), potentially leading to wide and uninformative predictions or problems with model convergence when not using such a shrinkage prior. All other parameters of the horseshoe priors were left at their default.

### Appendix 7: Prior predictive check

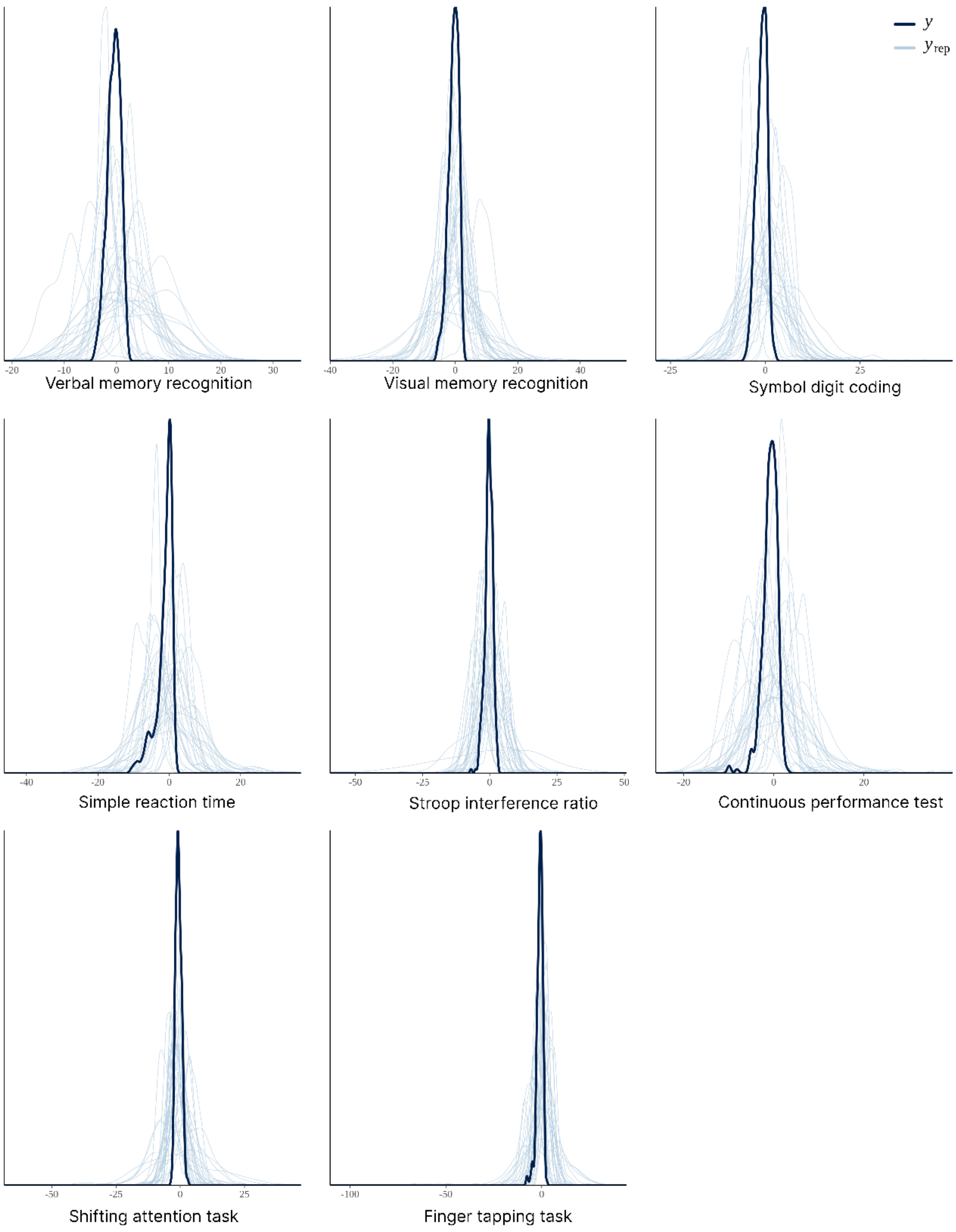


*Caption: Prior predictive check for the best performing model (model 4b), individually for each outcome measure. The dark blue line y represents the true output distribution. The light blue lines $y_{rep}$ represents the simulated data.*

### Appendix 8: Model parameters of the best-performing model (model 4b)

#### A: Fixed effects

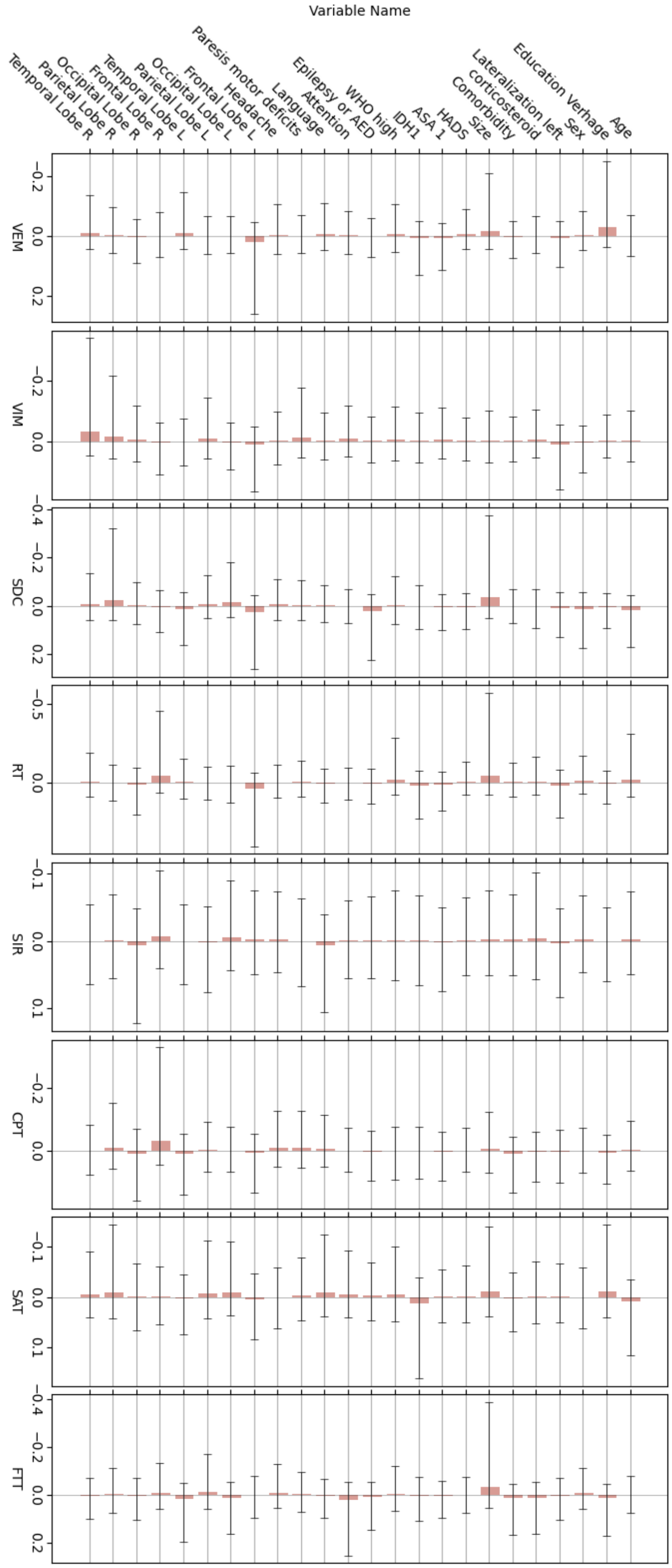

### B: Intercepts:

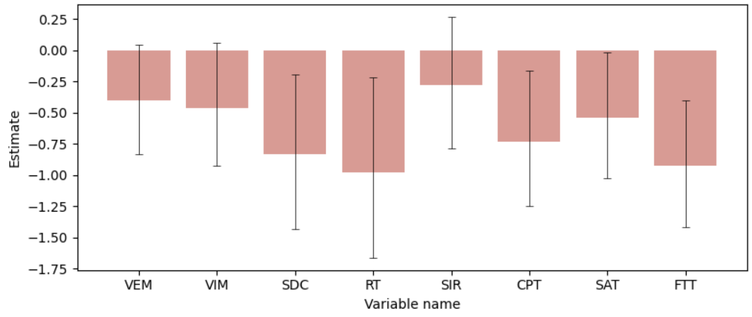


### C: Standard deviation of the likelyhood (sigma):

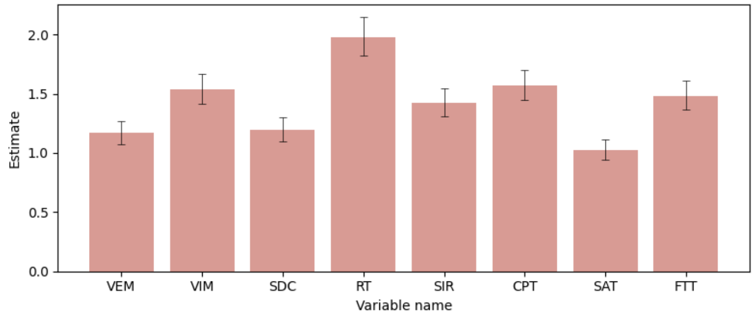


*Caption: Estimated coefficients, intercepts, and standard deviation of the likelyhood (mean, the pink bars) including their credibility interval (CI: 95%) for the model parameters of the best-performing model (model 4b). All random effects can be found as an online supplement. VEM: Verbal memory recognition, VIM: Visual memory recognition, SDC: Symbol digit coding task, RT: Simple reaction time, SI: Stroop interference ratio, CPT: Continuous performance task, SAT: Shifting attention task, FTT: Finger tapping task.*

### Appendix 9: Posterior predictive check

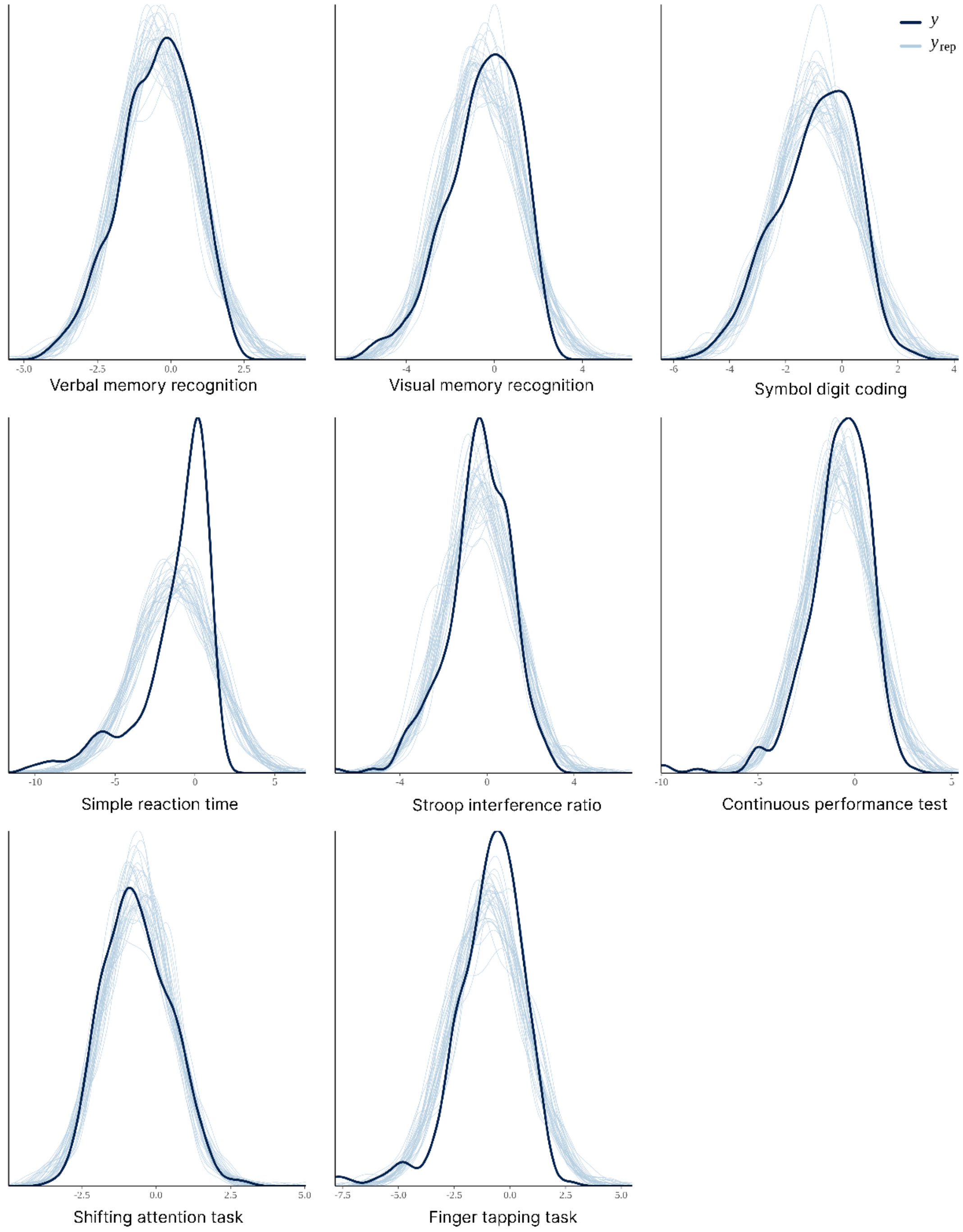


*Caption: Posterior predictive check for the best performing model (model 4b), individually for each outcome measure. The dark blue line y represents the true output distribution. The light blue lines $y_{rep}$ represents the simulated data.*
