## Supplementary material for "Modeling uncertainty in individual predictions of cognitive functioning for untreated glioma patients using Bayesian regression": Bayesian reporting checklist

### Key reporting points Bayesian Analysis Reporting Guidelines

1. Explain the model
   a. Data variables ✔
   b. Likelihood function and parameters ✔
   c. Prior distribution ✔
   d. Formal specification ✔
   e. Prior predictive check ✔
2. Report details of the computation
   a. Software ✔
   b. MCMC chain convergence ✔
   c. MCMC chain resolution ✔
3. Describe the posterior distribution
   a. Posterior predictive check ✔
   b. Summarize posterior of variables ✔
   c. BF and posterior model probabilities: NA.
4. Report decisions (if any) and their criteria: N.A
5. Report sensitivity analysis
   a. For broad priors ✔
   b. For informed priors: N.A.
   c. For default priors ✔
   d. BFs and model probabilities: N.A.
   e. Decisions: N.A.
6. Make it reproducible
   a. Software and installation ✔
   b. Software version details ✔
   c. Script and data ✔
   d. Readable for humans ✔
   e. All auxiliary files ✔
   f. Runs as posted ✔
   g. MCMC chains for time-intensive runs: N.A.
   h. Reproducible MCMC ✔
