## Supplementary material for "Modeling uncertainty in individual predictions of cognitive functioning for untreated glioma patients using Bayesian regression": R Code: Online supplement - code.R

```
library(brms)
library("stringr")
library("readxl")
library("MLmetrics")
library("glue")
library("matrixStats")
library("writexl")
library("purrr")
library("mice")
library("cmdstanr")
library("caret")
library("abind")
set.seed(23)


print(cmdstan_path())


path_to_data = "" #todo: add path to your data. See the example excel file attached as supplement
data <- read_excel(path_to_data)
colnames(data) = gsub(" ", "_", colnames(data))

# Settings for model fitting
do_kfold = TRUE
do_plot = FALSE
n_folds = 10 # for k-fold corss validation
n_mice = 30 #
library(future)
plan(multicore)
options(auto_write = TRUE)
options(mc.cores=parallel::detectCores())

# Output names
y_vars_nu = c('T0cVEM',
 'T0cVIM',
 'T0cSDC',
'T0cS1RT',
'T0cSIR32',
 'T0cCPT',
'T0cSAT',
 'T0cFTT')

y_vars = c('T0_c_VEM',
 'T0_c_VIM',
 'T0_c_SDC',
'T0_c_S1RT',
'T0_c_SIR32',
 'T0_c_CPT',
'T0_c_SAT',
 'T0_c_FTT')

# Predictor names
X_vars = c('age_T0',
 'education_Verhage',
 'sex',
 'lateralization_left',
 'T0_corticosteroid',
 'comorbidity',
 'size',
 'HADS',
 'ASA_1',
 'IDH1',
 'WHO_high',
 'epilepsy_or_AED',
 'Attention_executive_memory_behavioural_SB',
 'Language_SB',
 'Paresis_motor_deficits_SB',
 'Headache_nieuw_SB',
 'Frontal_Lobe_L',
 'Occipital_Lobe_L',
 'Parietal_Lobe_L',
 'Temporal_Lobe_L',
 'Frontal_Lobe_R',
 'Occipital_Lobe_R',
 'Parietal_Lobe_R',
 'Temporal_Lobe_R',
 'glioblastoma',
 'oligodendroglioma')
# Predictor names w/o tumor type for hierarchical models
X_vars_h = c('age_T0',
 'education_Verhage',
 'sex',
 'lateralization_left',
 'T0_corticosteroid',
 'comorbidity',
 'size',
 'HADS',
 'ASA_1',
 'IDH1',
 'WHO_high',
 'epilepsy_or_AED',
 'Attention_executive_memory_behavioural_SB',
 'Language_SB',
 'Paresis_motor_deficits_SB',
 'Headache_nieuw_SB',
 'Frontal_Lobe_L',
 'Occipital_Lobe_L',
 'Parietal_Lobe_L',
 'Temporal_Lobe_L',
 'Frontal_Lobe_R',
 'Occipital_Lobe_R',
 'Parietal_Lobe_R',
 'Temporal_Lobe_R'
)
# Create tumor categories
data[data$glioblastoma==TRUE, 'tumor_cat']=1
data[data$oligodendroglioma==TRUE, 'tumor_cat']=2
data[data$oligodendroglioma==FALSE & data$glioblastoma==FALSE, 'tumor_cat']=3

data['education_Verhage'] = as.factor(data$education_Verhage)
data['sex'] = as.factor(data$sex)
data['lateralization_left'] = as.factor(data$lateralization_left)
data['T0_corticosteroid'] = as.factor(data$T0_corticosteroid)
data['comorbidity'] = as.factor(data$comorbidity)
data['ASA_1'] = as.factor(data$ASA_1)
data['IDH1'] = as.factor(data$IDH1)
data['WHO_high'] = as.factor(data$WHO_high)
data['epilepsy_or_AED'] = as.factor(data$epilepsy_or_AED)
data['Attention_executive_memory_behavioural_SB'] = as.factor(data$Attention_executive_memory_behavioural_SB)
data['Language_SB'] = as.factor(data$Language_SB)
data['Paresis_motor_deficits_SB'] = as.factor(data$Paresis_motor_deficits_SB)
data['tumor_cat'] = as.factor(data$tumor_cat)


data_comp = data[complete.cases(data[c(y_vars)]),]
n <- nrow(data)
n_comp <- nrow(data_comp)
p <- length(y_vars)  # number of output features
# Generate balanced folds using caret
folds <- createFolds(1:n, k = n_folds, list = TRUE)
folds_comp <- createFolds(1:n_comp, k = n_folds, list = TRUE)


do_fit <- function(formula, priors, name, X_vars, y_vars_loc, complete_only, threads = 64, pp=TRUE) {
  print('Do fit')
  if (complete_only){
      data_loc = data_comp
  }
  else {
    data_loc = data
  }
  data_loc = data_loc[c(X_vars,y_vars_loc)]

  predictorMatrix <- matrix(0, ncol(data_loc), ncol(data_loc))
  predictorMatrix[match(c(X_vars), names(data_loc)), match(c(X_vars), names(data_loc))] <- 1
  init = mice(data_loc, maxit=0)
  method = init$method
  method[!names(method) %in% c(X_vars, 'tumor_cat')] <- ''

  imp_result = mice(data_loc, predictorMatrix=predictorMatrix, method=method, m = n_mice, print = TRUE)

  data_imputed = complete(imp_result, action='long')
  data_imputed[X_vars] = lapply(data_imputed[X_vars],as.numeric)
  preproc <- preProcess(data_imputed[c(X_vars)], method = c("center", "scale"))
  data_imputed[c(X_vars)] <- predict(preproc, data_imputed[c(X_vars)])
  data_imputed = split(data_imputed, data_imputed$.imp)

  backend = "cmdstanr"
  backend = "rstan"

  print('start brm_multiple')
  fit =
    brm_multiple(
      formula = formula,
      data = data_imputed,
      prior = priors,
      control = list(adapt_delta =.95),
      family = gaussian(),
      threads = 1,
      backend = backend,
      file = name,
      file_refit = 'always',
      save_model = name,
      silent = 0
    )

  fit_prior = NULL
  pp=FALSE
  if (pp==TRUE){
    print('Start pp')

    fit_prior =
      brm_multiple(
        formula = formula,
        data = data_imputed,
        prior = priors,
        control = list(adapt_delta =.95),
        family = gaussian(),
        threads = 1,
        backend = backend,
        file = paste0('prior', name),
        file_refit = 'always',
        save_model = paste0('prior', name),
        silent = 0,
        sample_prior = "only"
      )
    }
  print('End pp')

  print(fit)
  if (is.list(y_vars_loc)){
    for (one_y_var in y_vars_loc){
        one_y_var = gsub("_", "", one_y_var)
        loo_res_one = LOO(fit, newdata=na.omit(data_imputed[[1]]), resp = one_y_var)
        saveRDS(loo_res_one, file = paste('loo_one_', name, one_y_var, '.rds'))
    }
  }

  print('start loo')
  loo_res = LOO(fit, newdata=na.omit(data_imputed[[1]]))
  print('save loo')
  saveRDS(loo_res, file = paste('loo_', name, '.rds'))

  return(list(fit=fit, fitprior = fit_prior))
}
plot_distributions <- function(thefit, thefitprior, thefitname){
    if (do_plot) {
      n_params <- 20
      all_variables = names(thefit$fit@sim[[1]][[1]])
      total_params <- length(all_variables)
      total_plots <- ceiling(total_params / n_params)
      for (i in 1:total_plots) {
        print(i)
        start_idx <- (i - 1) * n_params + 1
        end_idx <- min(i * n_params, total_params)
        print(start_idx)
        print(end_idx)
        variables = all_variables[start_idx:end_idx]
        toplot = plot(thefit, N=length(variables) , variable = variables,
             combo = c("trace", "dens"), ask = FALSE, plot=FALSE)
        ggsave(paste0(thefitname, i, ".jpeg"), plot=toplot[[1]], width = 20, height = 35, bg = "white")
      }
      for (variable in y_vars_nu) {
        toplot = pp_check(thefitprior, resp = variable, ndraws=30)
        ggsave(paste0(thefitname, "_prior_", variable, ".jpeg"), plot=toplot, width = 15, height = 10, bg = "white")
        toplot = pp_check(thefit, resp = variable, ndraws=30)
        ggsave(paste0(thefitname, "_posterior_", variable, ".jpeg"), plot=toplot, width = 15, height = 10, bg = "white")
      }
   }
}
run_kfold <- function(formula, priors, name, X_vars, y_vars, complete_only, threads=64){
  # formula, priors, name, X_vars, to_predict, complete_only
  if (do_kfold) {
    print('Start kfold')

    if (complete_only){
        n_loc = n_comp
        folds_loc = folds_comp
        data_loc = data_comp
    }
    else {
          n_loc = n
          folds_loc = folds
          data_loc = data
    }
    data_loc = data_loc[c(X_vars,y_vars)]

    backend = "cmdstanr"
    #backend = "rstan"

    # Initialize variables to store results
    all_predsmv <- matrix(NA, n_loc, length(y_vars))  # to store all predictions

    for(i in 1:n_folds) {
      print('Start a fold')
      test_idx <- folds_loc[[i]]
      train_idx <- setdiff(1:n, test_idx)

      train_data <- data_loc[train_idx, , drop = FALSE]
      test_data <- data_loc[test_idx, , drop = FALSE]

      predictorMatrix <- matrix(0, ncol(data_loc), ncol(data_loc))
      predictorMatrix[match(c(X_vars), names(data_loc)), match(c(X_vars), names(data_loc))] <- 1
      init = mice(data_loc, maxit=0)
      method = init$method
      method[!names(method) %in% c(X_vars, 'tumor_cat')] <- ''

      imp_train <- mice(train_data, predictorMatrix=predictorMatrix, method=method, m = n_mice, print = TRUE)
      imp_test <- mice.mids(imp_train, newdata = test_data)

      compl_train = complete(imp_train, action='long')
      compl_train[X_vars] = lapply(compl_train[X_vars],as.numeric)
      preproc <- preProcess(train_data[c(X_vars)], method = c("center", "scale"))
      compl_train[c(X_vars)] <- predict(preproc, compl_train[c(X_vars)])
      compl_train = split(compl_train, compl_train$.imp)
      compl_test = complete(imp_test, action='long')
      compl_test[X_vars] = lapply(compl_test[X_vars],as.numeric)
      compl_test[c(X_vars)] <- predict(preproc, compl_test[c(X_vars)])
      compl_test = split(compl_test, compl_test$.imp)

      print('before brm_multiple in kfold')
      # Fit the brms model on the training data
      fitted_model =
        brm_multiple(
          formula = formula,
          data = compl_train,
          prior = priors,
          control = list(adapt_delta =.95),
          family = gaussian(),
          #future = TRUE,
          threads = 1,
          backend = backend,
          file = paste0(name, 'kfold', i),
          file_refit = 'always',
          # save_model = paste0('fit_mv', i),
          silent = 0
        )

      print('Creating predictions')
      # Generate predictions using the fitted brms model. Average predictions of the different imputed datasets
      for (j_mice in 1:n_mice){
        comp_test_loc = compl_test[[j_mice]]
        if (j_mice == 1){
          y_pred <- posterior_predict(fitted_model, newdata = comp_test_loc)
        }
        else {
          y_pred = abind(y_pred, posterior_predict(fitted_model, newdata = comp_test_loc), along = 1)
        }
      }

      print("Saving draws")
      print(dim(y_pred))
      savedraws = list(test_idx, y_pred)
      saveRDS(savedraws, file = paste('savedraws', name, '_', i, '.rds'))
      y_pred <- colMeans(y_pred, na.rm=TRUE) # Averaging over all posterior samples resulting from the different datasets
      all_predsmv[test_idx, ] <- y_pred

    }
    # saveRDS(all_draws, file = paste('All_drawsmv', name, '.rds'))

    print('Calc R2')
    if (complete_only == FALSE){
      R2_mv <- sapply(1:length(y_vars), function(i) {
        print('R2 normal')
        col1 <- all_predsmv[, i, drop=FALSE]
        col2 <- data_loc[[y_vars[i]]]
        print(dim(data_loc))
        print(length(data_loc))
        valid_rows <- complete.cases(col1, col2) # Only y var of intrest needs to be complete
        correlation <- cor(col1[valid_rows], col2[valid_rows])
        correlation^2
      })
      print(R2_mv)
      saveRDS(R2_mv, file = paste('R2_', name, '.rds'))
    }

    R2_mv_comp <- sapply(1:length(y_vars), function(i) {
      print('R2 complete')
      col1 <- all_predsmv[, i, drop=FALSE]
      col2 <- data_loc[[y_vars[i]]]
      print(dim(data_loc))
      print(length(data_loc))
      valid_rows <- complete.cases(col1, data_loc[, y_vars]) # All y_vars need to be complete
      correlation <- cor(col1[valid_rows], col2[valid_rows])
      correlation^2
    })
    print(R2_mv_comp)
    saveRDS(R2_mv_comp, file = paste('R2_comp_', name, '.rds'))

  }
}


print('------------------------------------------------')
print('New model')
print('Frequentist LM')
if (FALSE) {

  predictors = paste(X_vars, collapse ='+')
  if (do_kfold) {

    all_predsmv <- matrix(NA, n, p)  # to store all predictions

    for (to_predict_i in 1:length(y_vars)){
      y_var = y_vars[to_predict_i]
      # Initialize variables to store results

      for(i_fold in 1:n_folds) {
        test_idx <- folds[[i_fold]]
        train_idx <- setdiff(1:n, test_idx)

        data_loc = data[c(X_vars,y_vars)]
        train_data <- data_loc[train_idx, , drop = FALSE]
        test_data <- data_loc[test_idx, , drop = FALSE]

        predictorMatrix <- matrix(0, ncol(data_loc), ncol(data_loc))
        predictorMatrix[match(c(X_vars), names(data_loc)), match(c(X_vars), names(data))] <- 1
        init = mice(data_loc, maxit=0)
        method = init$method
        method[!names(method) %in% c(X_vars, 'tumor_cat')] <- ''

        imp_train <- mice(train_data, predictorMatrix=predictorMatrix, method=method, m = n_mice, print = TRUE)
        imp_test <- mice.mids(imp_train, newdata = test_data)

        compl_train = complete(imp_train, action='long')
        preproc <- preProcess(train_data[c(X_vars)], method = c("center", "scale"))
        compl_train[c(X_vars)] <- predict(preproc, compl_train[c(X_vars)])
        compl_train = split(compl_train, compl_train$.imp)
        compl_test = complete(imp_test, action='long')
        compl_test[c(X_vars)] <- predict(preproc, compl_test[c(X_vars)])
        compl_test = split(compl_test, compl_test$.imp)

        formula = glue("{y_var} ~ {predictors}")

        # regression = lm(formula, data=train_data, na.action='na.omit')
        # y_pred = predict(regression, compl_test)

        regression = lm.mids(formula, data=imp_train, na.action='na.omit')

        for (j_mice in 1:n_mice){
          comp_test_loc = compl_test[[j_mice]]

          # https://github.com/amices/mice/issues/82
          predm = lapply(getfit(regression), predict, newdata=comp_test_loc, se.fit = TRUE)
          Q <- sapply(predm, `[[`, "fit")
          U <- sapply(predm, `[[`, "se.fit")^2
          dfcom <- predm[[1]]$df
          pred <- matrix(NA, nrow = nrow(Q), ncol = 3,
                     dimnames = list(NULL, c("fit", "se.fit", "df")))
          for(k in 1:nrow(Q)) {
            pi <- pool.scalar(Q[k, ], U[k, ], n = dfcom + 1)
            pred[k, 1] <- pi[["qbar"]]
            pred[k, 2] <- sqrt(pi[["t"]])
            pred[k, 3] <- pi[["df"]]
          }

          if (j_mice == 1){
            y_pred <- pred[, 1]
          }
          else {
            y_pred = Map(c, y_pred, pred[, 1])
          }
        }

        # y_pred = do.call("cbind",y_pred) #todo: CHANGE for not for nmice = 1
        y_pred = do.call("cbind",list(y_pred)) #todo: CHANGE for nmice = 1
        y_pred <- colMeans(y_pred, na.rm = TRUE)
        print(y_pred)
        print(length(y_pred))
        all_predsmv[test_idx, to_predict_i] <- y_pred
      }
    }

    complete_only = FALSE
    if (complete_only == FALSE){
      R2_mv <- sapply(1:p, function(i) {
        print('R2 normal')
        col1 <- all_predsmv[, i, drop=FALSE]
        col2 <- data[[y_vars[i]]]
        print(dim(data))
        print(length(data))
        valid_rows <- complete.cases(col1, col2) # Only y var of intrest needs to be complete
        correlation <- cor(col1[valid_rows], col2[valid_rows])
        correlation^2
      })
      print(R2_mv)
      #saveRDS(R2_mv, file = 'R2_fqlm_nmice1.rds') #todo: CHANGE BACK
      saveRDS(R2_mv, file = 'R2_fqlm_reallynmice1.rds') #todo: CHANGE BACK
    }

    R2_mv_comp <- sapply(1:p, function(i) {
      print('R2 complete')
      col1 <- all_predsmv[, i, drop=FALSE]
      col2 <- data[[y_vars[i]]]
      print(dim(data))
      print(length(data))
      valid_rows <- complete.cases(col1, data[, y_vars]) # All y_vars need to be complete
      correlation <- cor(col1[valid_rows], col2[valid_rows])
      correlation^2
    })
    print(R2_mv_comp)
    saveRDS(R2_mv_comp, file = 'R2_comp_fqlm_nmice1.rds') #todo: CHANGE BACK
    #saveRDS(R2_mv_comp, file = 'R2_comp_fqlm.rds') #todo: CHANGE BACK

  }
}
# Server 1
print('------------------------------------------------')
print('New model')
print('Multivariate')
if (FALSE) {

  predictors = paste(X_vars, collapse ='+')
  output_vars = paste(y_vars, collapse = ', ')
  formula = bf(glue("mvbind({output_vars}) | mi() ~ {predictors}"))

  priors = c(
        reduce(map(y_vars_nu, ~ set_prior("horseshoe(1, par_ratio  = 0.8)", class="b", resp =.x)), `+`) ,
        reduce(map(y_vars_nu, ~ set_prior("normal(0, 3)", class = "sigma", resp = .x)),`+`) ,
        reduce(map(y_vars_nu, ~ set_prior("normal(0, 3)", class = "Intercept", resp = .x)),`+`) , # Average = -.70
        prior(lkj(6), class = rescor)
  )

  name = 'multivariate'
  rescor = TRUE
  complete_only = FALSE
  formula = formula + set_rescor(rescor)

  fits = do_fit(formula, priors, name, X_vars, y_vars, complete_only)
  fit = fits$fit
  fitprior = fits$fitprior
  plot_distributions(fit, fitprior, name)
  run_kfold(formula, priors, name, X_vars, y_vars, complete_only)
}

print('------------------------------------------------')
print('New model')
print('Multivariate no residual correlations')
if (FALSE) {

  predictors = paste(X_vars, collapse ='+')
  output_vars = paste(y_vars, collapse = ', ')
  formula = bf(glue("mvbind({output_vars}) | mi() ~ {predictors}"))

  priors = c(
        reduce(map(y_vars_nu, ~ set_prior("horseshoe(1, par_ratio  = 0.8)", class="b", resp =.x)), `+`) ,
        reduce(map(y_vars_nu, ~ set_prior("normal(0, 3)", class = "sigma", resp = .x)),`+`) ,
        reduce(map(y_vars_nu, ~ set_prior("normal(0, 3)", class = "Intercept", resp = .x)),`+`)  # Average = -.70
  )

  name = "Multivariate no residual correlations"
  rescor = FALSE
  complete_only = FALSE
  formula = formula + set_rescor(rescor)

  fits = do_fit(formula, priors, name, X_vars, y_vars, complete_only)
  fit = fits$fit
  fitprior = fits$fitprior
  plot_distributions(fit, fitprior, name)
  run_kfold(formula, priors, name, X_vars, y_vars, complete_only)
}

print('------------------------------------------------')
print('New model')
print('No pooling')
if (FALSE) {


  predictors = paste(X_vars_h, collapse ='+')
  output_vars = paste(y_vars, collapse = ', ')
  formula = bf(glue("mvbind({output_vars}) | mi() ~ ({predictors}) * as.factor(tumor_cat)"))

  priors = c(
    reduce(map(y_vars_nu, ~ set_prior("horseshoe(1, par_ratio  = 0.8)", class="b", resp =.x)), `+`) ,
    reduce(map(y_vars_nu,~ set_prior("normal(0, 3)", class = "sigma", resp = .x)),`+`) ,
    reduce(map(y_vars_nu,~ set_prior("normal(0, 3)", class = "Intercept", resp = .x)),`+`),
    prior(lkj(6), class = rescor))

  name = 'No pooling'
  rescor = TRUE
  complete_only = FALSE
  formula = formula + set_rescor(rescor)

  fits = do_fit(formula, priors, name, c(X_vars_h, 'tumor_cat'), y_vars, complete_only)
  fit = fits$fit
  fitprior = fits$fitprior
  plot_distributions(fit, fitprior, name)
  run_kfold(formula, priors, name, c(X_vars_h, 'tumor_cat'), y_vars, complete_only)
}

print('------------------------------------------------')
print('New model')
print('No pooling, no residual correlations')
if (FALSE) {

  predictors = paste(X_vars_h, collapse ='+')
  output_vars = paste(y_vars, collapse = ', ')
  formula = bf(glue("mvbind({output_vars}) | mi() ~ ({predictors}) * as.factor(tumor_cat)"))

  priors = c(
    reduce(map(y_vars_nu, ~ set_prior("horseshoe(1, par_ratio  = 0.8)", class="b", resp =.x)), `+`) ,
    reduce(map(y_vars_nu,~ set_prior("normal(0, 3)", class = "sigma", resp = .x)),`+`) ,
    reduce(map(y_vars_nu,~ set_prior("normal(0, 3)", class = "Intercept", resp = .x)),`+`))

  name = 'No pooling no residual correlations'
  rescor = FALSE
  complete_only = FALSE
  formula = formula + set_rescor(rescor)

  fits = do_fit(formula, priors, name, c(X_vars_h, 'tumor_cat'), y_vars, complete_only)
  fit = fits$fit
  fitprior = fits$fitprior
  plot_distributions(fit, fitprior, name)
  run_kfold(formula, priors, name, c(X_vars_h, 'tumor_cat'), y_vars, complete_only)
}

print('------------------------------------------------')
print('New model')
print('Partial pooling') #1
# todo: always run this one seperately or at the end as threading does not work
if (FALSE) {


  # Create formula
  predictors = paste(X_vars_h, collapse ='+')
  output_vars = paste(y_vars, collapse = ', ')
  formula = bf(glue("mvbind({output_vars}) | mi() ~ {predictors}  + (1 + {predictors} | tumor_cat)"))
  # Instead of | you may use || in grouping terms to prevent correlations from being modeled.

  priors = c(
    reduce(map(y_vars_nu, ~ set_prior("horseshoe(1, par_ratio  = 0.8)", class="b", resp =.x)), `+`) ,
    reduce(map(y_vars_nu,~ set_prior("normal(0, 3)", class = "sigma", resp = .x)),`+`) ,
    reduce(map(y_vars_nu,~ set_prior("normal(0, 3)", class = "Intercept", resp = .x)),`+`),
    reduce(map(y_vars_nu,~ set_prior("normal(0, 0.5)", class = "sd", resp = .x)),`+`) ,
    prior(lkj(6), class = rescor)
  )

  name = 'Partial pooling2'
  name = 'Partial pooling2 JUSTATESTDONTSAVE'
  rescor = TRUE
  complete_only = FALSE
  formula = formula + set_rescor(rescor)

  fits = do_fit(formula, priors, name, c(X_vars_h, 'tumor_cat'), y_vars, complete_only, NULL)
  fit = fits$fit
  fitprior = fits$fitprior
  #plot_distributions(fit, fitprior, name)
  #run_kfold(formula, priors, name, c(X_vars_h, 'tumor_cat'), y_vars, complete_only, NULL)
}

print('------------------------------------------------')
print('New model') #2
print('Partial pooling, no residual correlations')
# todo: always run this one seperately or at the end as threading does not work
if (FALSE) {


  # Create formula
  predictors = paste(X_vars_h, collapse ='+')
  output_vars = paste(y_vars, collapse = ', ')
  formula = bf(glue("mvbind({output_vars}) | mi() ~ {predictors}  + (1 + {predictors} | tumor_cat)"))

  priors = c(
    reduce(map(y_vars_nu, ~ set_prior("horseshoe(1, par_ratio  = 0.8)", class="b", resp =.x)), `+`) ,
    reduce(map(y_vars_nu,~ set_prior("normal(0, 3)", class = "sigma", resp = .x)),`+`) ,
    reduce(map(y_vars_nu,~ set_prior("normal(0, 3)", class = "Intercept", resp = .x)),`+`),
    reduce(map(y_vars_nu,~ set_prior("normal(0, 0.5)", class = "sd", resp = .x)),`+`) )

  name = 'partial pooling no rescor fix'
  rescor = FALSE
  complete_only = FALSE
  formula = formula + set_rescor(rescor)

  fits = do_fit(formula, priors, name, c(X_vars_h, 'tumor_cat'), y_vars, complete_only, NULL, FALSE)
  # fit = fits$fit
  # fitprior = fits$fitprior
  # plot_distributions(fit, fitprior, name)
  # run_kfold(formula, priors, name, c(X_vars_h, 'tumor_cat'), y_vars, complete_only, NULL)
}


print('------------------------------------------------')
print('Sensitivity test')
print('------------------------------------------------')
print('------------------------------------------------')
print('------------------------------------------------')
print('------------------------------------------------')
print('------------------------------------------------')
print('New model')
print('Partial pooling norescor no hs') #3
if (FALSE) {

  # Create formula
  predictors = paste(X_vars_h, collapse ='+')
  output_vars = paste(y_vars, collapse = ', ')
  formula = bf(glue("mvbind({output_vars}) | mi() ~ {predictors}  + (1 + {predictors} | tumor_cat)"))
  # Instead of | you may use || in grouping terms to prevent correlations from being modeled.

  priors = c(
    # reduce(map(y_vars_nu, ~ set_prior("horseshoe(1, par_ratio  = 0.8)", class="b", resp =.x)), `+`) ,
    reduce(map(y_vars_nu,~ set_prior("normal(0, 3)", class = "sigma", resp = .x)),`+`) ,
    reduce(map(y_vars_nu,~ set_prior("normal(0, 3)", class = "Intercept", resp = .x)),`+`),
    reduce(map(y_vars_nu,~ set_prior("normal(0, 0.5)", class = "sd", resp = .x)),`+`) )
  #priors = NULL

  name = 'Partial pooling no rescor no horseshoe'
  rescor = FALSE
  complete_only = FALSE
  formula = formula + set_rescor(rescor)

  fits = do_fit(formula, priors, name, c(X_vars_h, 'tumor_cat'), y_vars, complete_only, NULL, pp=FALSE)
  fit = fits$fit
  fitprior = fits$fitprior
  plot_distributions(fit, fitprior, name)
  run_kfold(formula, priors, name, c(X_vars_h, 'tumor_cat'), y_vars, complete_only, NULL)
}
print('------------------------------------------------')
print('New model')
print('Partial pooling norescor only hs') #4
if (FALSE) {

  # Create formula
  predictors = paste(X_vars_h, collapse ='+')
  output_vars = paste(y_vars, collapse = ', ')
  formula = bf(glue("mvbind({output_vars}) | mi() ~ {predictors}  + (1 + {predictors} | tumor_cat)"))
  # Instead of | you may use || in grouping terms to prevent correlations from being modeled.

  priors = c(
    reduce(map(y_vars_nu, ~ set_prior("horseshoe(1, par_ratio  = 0.8)", class="b", resp =.x)), `+`)
    # reduce(map(y_vars_nu,~ set_prior("normal(0, 3)", class = "sigma", resp = .x)),`+`) ,
    # reduce(map(y_vars_nu,~ set_prior("normal(0, 3)", class = "Intercept", resp = .x)),`+`),
    # reduce(map(y_vars_nu,~ set_prior("normal(0, 0.5)", class = "sd", resp = .x)),`+`) ,
    # prior(lkj(6), class = rescor)
  )
  #priors = NULL

  name = 'Partial pooling no rescor only horseshoe'
  rescor = FALSE
  complete_only = FALSE
  formula = formula + set_rescor(rescor)

  fits = do_fit(formula, priors, name, c(X_vars_h, 'tumor_cat'), y_vars, complete_only, NULL, pp=FALSE)
  fit = fits$fit
  fitprior = fits$fitprior
  plot_distributions(fit, fitprior, name)
  run_kfold(formula, priors, name, c(X_vars_h, 'tumor_cat'), y_vars, complete_only, NULL)
}
print('------------------------------------------------')
print('New model')
print('Partial pooling no rescor default priors') #5
if (FALSE) {

  # Create formula
  predictors = paste(X_vars_h, collapse ='+')
  output_vars = paste(y_vars, collapse = ', ')
  formula = bf(glue("mvbind({output_vars}) | mi() ~ {predictors}  + (1 + {predictors} | tumor_cat)"))
  # Instead of | you may use || in grouping terms to prevent correlations from being modeled.

  priors = c(
    reduce(map(y_vars_nu, ~ set_prior("horseshoe(1, par_ratio  = 0.8)", class="b", resp =.x)), `+`) ,
    reduce(map(y_vars_nu,~ set_prior("normal(0, 3)", class = "sigma", resp = .x)),`+`) ,
    reduce(map(y_vars_nu,~ set_prior("normal(0, 3)", class = "Intercept", resp = .x)),`+`),
    reduce(map(y_vars_nu,~ set_prior("normal(0, 0.5)", class = "sd", resp = .x)),`+`) )
  priors = NULL

  name = 'Partial pooling default priors'
  rescor = FALSE
  complete_only = FALSE
  formula = formula + set_rescor(rescor)

  # fits = do_fit(formula, priors, name, c(X_vars_h, 'tumor_cat'), y_vars, complete_only, NULL, pp=FALSE)
  # fit = fits$fit
  # fitprior = fits$fitprior
  # plot_distributions(fit, fitprior, name)
  run_kfold(formula, priors, name, c(X_vars_h, 'tumor_cat'), y_vars, complete_only, NULL)
}
print('------------------------------------------------')
print('------------------------------------------------')
print('------------------------------------------------')
print('------------------------------------------------')
print('------------------------------------------------')
print('New model')
print('Multivariate no hs')
if (FALSE) {

  predictors = paste(X_vars, collapse ='+')
  output_vars = paste(y_vars, collapse = ', ')
  formula = bf(glue("mvbind({output_vars}) | mi() ~ {predictors}"))

  priors = c(
        #reduce(map(y_vars_nu, ~ set_prior("horseshoe(1, par_ratio  = 0.8)", class="b", resp =.x)), `+`) ,
        reduce(map(y_vars_nu, ~ set_prior("normal(0, 3)", class = "sigma", resp = .x)),`+`) ,
        reduce(map(y_vars_nu, ~ set_prior("normal(0, 3)", class = "Intercept", resp = .x)),`+`)  # Average = -.70
  )

  name = "Multivariate no hs"
  rescor = FALSE
  complete_only = FALSE
  formula = formula + set_rescor(rescor)

  # fits = do_fit(formula, priors, name, X_vars, y_vars, complete_only)
  # fit = fits$fit
  # fitprior = fits$fitprior
  # plot_distributions(fit, fitprior, name)
  run_kfold(formula, priors, name, X_vars, y_vars, complete_only)
}
print('New model')
print('Multivariate only hs')
if (FALSE) {

  predictors = paste(X_vars, collapse ='+')
  output_vars = paste(y_vars, collapse = ', ')
  formula = bf(glue("mvbind({output_vars}) | mi() ~ {predictors}"))

  priors = c(
        reduce(map(y_vars_nu, ~ set_prior("horseshoe(1, par_ratio  = 0.8)", class="b", resp =.x)), `+`)# ,
        #reduce(map(y_vars_nu, ~ set_prior("normal(0, 3)", class = "sigma", resp = .x)),`+`) ,
        #reduce(map(y_vars_nu, ~ set_prior("normal(0, 3)", class = "Intercept", resp = .x)),`+`)  # Average = -.70
  )

  name = "Multivariate only hs"
  rescor = FALSE
  complete_only = FALSE
  formula = formula + set_rescor(rescor)

  # fits = do_fit(formula, priors, name, X_vars, y_vars, complete_only)
  # fit = fits$fit
  # fitprior = fits$fitprior
  # plot_distributions(fit, fitprior, name)
  run_kfold(formula, priors, name, X_vars, y_vars, complete_only)
}
print('New model')
print('Multivariate default priors')
if (FALSE) {

  predictors = paste(X_vars, collapse ='+')
  output_vars = paste(y_vars, collapse = ', ')
  formula = bf(glue("mvbind({output_vars}) | mi() ~ {predictors}"))

  priors = c(
        reduce(map(y_vars_nu, ~ set_prior("horseshoe(1, par_ratio  = 0.8)", class="b", resp =.x)), `+`) ,
        reduce(map(y_vars_nu, ~ set_prior("normal(0, 3)", class = "sigma", resp = .x)),`+`) ,
        reduce(map(y_vars_nu, ~ set_prior("normal(0, 3)", class = "Intercept", resp = .x)),`+`)  # Average = -.70
  )
  priors = NULL

  name = "Multivariate default priors"
  rescor = FALSE
  complete_only = FALSE
  formula = formula + set_rescor(rescor)

  # fits = do_fit(formula, priors, name, X_vars, y_vars, complete_only)
  # fit = fits$fit
  # fitprior = fits$fitprior
  # plot_distributions(fit, fitprior, name)
  run_kfold(formula, priors, name, X_vars, y_vars, complete_only)
}
print('New model')
print('Multivariate default priors N_MICE1')
if (FALSE) {

  predictors = paste(X_vars, collapse ='+')
  output_vars = paste(y_vars, collapse = ', ')
  formula = bf(glue("mvbind({output_vars}) | mi() ~ {predictors}"))

  priors = c(
        reduce(map(y_vars_nu, ~ set_prior("horseshoe(1, par_ratio  = 0.8)", class="b", resp =.x)), `+`) ,
        reduce(map(y_vars_nu, ~ set_prior("normal(0, 3)", class = "sigma", resp = .x)),`+`) ,
        reduce(map(y_vars_nu, ~ set_prior("normal(0, 3)", class = "Intercept", resp = .x)),`+`)  # Average = -.70
  )
  priors = NULL

  name = "Multivariate default priors onemice"
  rescor = FALSE
  complete_only = FALSE
  formula = formula + set_rescor(rescor)

  n_mice_orig = n_mice
  n_mice = 1
  # fits = do_fit(formula, priors, name, X_vars, y_vars, complete_only)
  # fit = fits$fit
  # fitprior = fits$fitprior
  # plot_distributions(fit, fitprior, name)
  run_kfold(formula, priors, name, X_vars, y_vars, complete_only)
  m_mice = n_mice_orig
}
```
